# Genetic structure of major depression symptoms across clinical and community cohorts

**DOI:** 10.1101/2023.07.05.23292214

**Authors:** Mark J Adams, Jackson G Thorp, Bradley S Jermy, Alex S F Kwong, Kadri Kõiv, Andrew D Grotzinger, Michel G Nivard, Sally Marshall, Yuri Milaneschi, Bernhard T Baune, Bertram Müller-Myhsok, Brenda WJH Penninx, Dorret I Boomsma, Douglas F Levinson, Gerome Breen, Giorgio Pistis, Hans J Grabe, Henning Tiemeier, Klaus Berger, Marcella Rietschel, Patrik K Magnusson, Rudolf Uher, Steven P Hamilton, Susanne Lucae, Kelli Lehto, Qingqin S Li, Enda M Byrne, Ian B Hickie, Nicholas G Martin, Sarah E Medland, Naomi R Wray, Elliot M Tucker-Drob, Estonian Biobank Research Team, Major Depressive Disorder Working Group of the Psychiatric Genomics Consortium, Cathryn M Lewis, Andrew M McIntosh, Eske M Derks

## Abstract

Diagnostic criteria for major depressive disorder allow for heterogeneous symptom profiles but genetic analysis of major depressive symptoms has the potential to identify clinical and aetiological subtypes. There are several challenges to integrating symptom data from genetically-informative cohorts, such as sample size differences between clinical and community cohorts and various patterns of missing data. We conducted genome-wide association studies of major depressive symptoms in three clinical cohorts that were enriched for affected participants (Psychiatric Genomics Consortium, Australian Genetics of Depression Study, Generation Scotland) and three community cohorts (Avon Longitudinal Study of Parents and Children, Estonian Biobank, and UK Biobank). We fit a series of confirmatory factor models with factors that accounted for how symptom data was sampled and then compared alternative models with different symptom factors. The best fitting model had a distinct factor for *Appetite/Weight* symptoms and an additional measurement factor that accounted for missing data patterns in the community cohorts (use of Depression and Anhedonia as gating symptoms). The results show the importance of assessing the directionality of symptoms (such as hypersomnia versus insomnia) and of accounting for study and measurement design when meta-analysing genetic association data.

## Introduction

Major depressive disorder (MDD) is a mood disorder characterized by low mood, loss of interest or pleasure (anhedonia), irritable affect, biological symptoms (psychomotor agitation/slowing, altered sleep patterns, changes in appetite or weight), negative thought content, and associated loss of function. To qualify for a diagnosis of major depression, the standard diagnostic classification systems (American Psychiatric Association, 2000, 2013; World Health Organization, 1992) require one of two cardinal symptoms plus at least four other symptoms to be present. Although conceptualized as a single disorder, the diagnostic criterion for MDD can be met with any combination of these other symptoms. For the DSM-5, this entails that there are 227 symptom profiles that would lead to a diagnosis of major depression (Zimmerman et al., 2015). When considering all potential symptom states (such as increasing versus decreasing appetite) the number of possible symptom profiles blooms into the thousands (Fried & Nesse, 2015a).

A single categorical phenotype---that might mask a multitude of separate disorder types--- stymies the testing of correlates and treatments. Network analysis has shown that MDD symptoms are not all equally related to each other (Borsboom & Cramer, 2013) and latent class analysis has been used to identify several MDD subtypes with differing patterns of symptoms and differential association with demographic, psychological, and physical health factors (Lamers et al., 2010). However, the potential concealed heterogeneity within the MDD diagnosis does have an upper bound: only around one quarter of the potential symptom profiles are actually observed (Fried & Nesse, 2015a; Zimmerman et al., 2015). This suggests there is both regularity and variation in symptom presentation.

Analysing individual symptoms is one way to unwrap the heterogeneity of MDD (Cai et al., 2020; Fried & Nesse, 2015b). Phenotypic studies have derived and tested factor structures of MDD symptoms (Elhai et al., 2012; Krause et al., 2008, 2010) and twin models have been used to separate genetic from environmental sources of symptom covariance (Kendler et al., 2013). These models grouped symptoms together in two or three factors, which broadly contrast psychological versus somatic symptoms. The primary difference among the proposed two factor structures is whether psychological symptoms including anhedonia and concentration problems group with the cognitive/affective symptoms or with the somatic symptoms. Three factor models have instead posited splitting the psychological symptoms into affective and cognitive components. Clinical subtypes are also part of diagnostic criteria and these have been used to classify depression profiles that are differentially associated with specific clinical, behavioural and biological correlates (Milaneschi et al., 2020; Penninx et al., 2013).

More recently, genetic studies of depressive symptoms have updated the findings from twin models using data from genome-wide association studies (GWAS). A confirmatory factor analysis of genetic covariance estimates obtained from GWAS results on current depressive symptoms showed that a psychological and somatic factor had the best fit to the data (Thorp et al., 2020). The detection of genetic correlates specific to each symptom implies that symptoms may have differing genetic causes and consequences, even if the symptoms themselves are highly genetically correlated.

Understanding the genetic architecture of MDD symptoms is complicated by symptom ascertainment. In clinical samples, symptom data is often only available on affected participants, and is thus conditioned on having been diagnosed with depression. Conditioning data presence on a diagnosis can induce downward bias in correlations amongst the symptoms comprising that diagnosis. However, when symptom data is missing in controls, imputing the absence of the symptom in controls and including them in the analysis has the potential to recapitulate the signal from a case/control analysis rather than reveal genetic variance that is unique to each symptom. In community cohorts, participants are typically screened for the presence of cardinal symptoms (depressed mood and anhedonia) and only participants who report at least one cardinal symptom are assessed for other symptoms of depression, which also leads to high levels of missing symptom data in these cohorts. Moreover, because community samples often contain symptom but not diagnostic information, many GWAS purporting to investigate MDD may actually be better characterized as investigating a broader dysphoria continuum rather than MDD specifically (Flint, 2023). Because community cohorts tend to have a larger sample size than clinical cohorts, meta-analysing all data together therefore has the potential to dilute information on case subtypes.

In this study we sought to uncover the genetic structure of depression symptoms while accounting for how samples were recruited and how symptoms were assessed. We did this by conducting GWAS of individual symptoms of depression, testing factor models to investigate genetic heterogeneity as a function of sample ascertainment (Clinical vs Community) and measurement (with or without screening based on cardinal/gating symptoms). Finally, we assessed the validity of the identified latent factors of depression by estimating genetic correlations with external traits.

Specifically, we conducted GWAS of symptom data in six cohorts and meta-analysed them in groups based on sample ascertainment. The first group (the “Clinical” cohorts) consisted of clinical cases from the Psychiatric Genomics Consortium MDD cohorts, participants from the Australian Genetics of Depression study who were recruited based on depression diagnosis, and participants from Generation Scotland who met DSM criteria for depression. The second group (the “Community” cohorts) consisted of the Avon Longitudinal Study of Parents and Children, Estonian Biobank, and UK Biobank, and thus contained data on participants who were not recruited with respect to depression status. Using the two sets of meta-analysed symptom GWASs, we first constructed and tested factor models that accounted for how the samples were recruited (Clinical versus Community) and how symptoms were assessed (such as gating symptoms in the Community cohorts). After understanding the measurements structure of the symptom GWASs, we then compared alternative factor models for the symptoms based on previous literature and diagnostic specifiers for depressive disorders. Using the best fitting overall model, we tested for shared and specific genetic correlates with other psychiatric, behavioral, and metabolic phenotypes that have known genetic links to MDD.

## Methods

### Samples and symptom measures

We analysed depression symptom data in six studies: the Psychiatric Genomics Consortium, the Australian Genetics of Depression Study, Generation Scotland, the Avon Longitudinal Study of Parents and Children, Estonian Biobank, and UK Biobank. Table 1 describes the number of participants with and without each symptom for each grouping of studies that were analysed. See Supplementary Material for information on genotyping and imputation.

**Table 1.** Sample size counts and sample prevalences of presence and absence of each symptom for participants used in the genetic analyses. Meta-analysis of Clinical (PGC, AGDS, GenScot), Community (ALSPAC, EstBB, UKB-MHQ) and UKB Touchscreen.

|  |  | **Cohort**<br>**N Symptom Present : Absent**<br>**Sample Prevalence** |  |  |
| --- | --- | --- | --- | --- |
| **Symptom** | **Abbr.** | <u>Clinical cohorts meta</u> | <u>Community cohorts meta</u> | <u>UKB Touchscreen</u> |
| 1. Depressed mood | Dep | 21681 : 1748<br>93% | 107956 : 99480<br>52% | 71964 : 57130<br>56% |
| 2. Anhedonia | Anh | 24732 : 2801<br>90% | 181113 : 126167<br>39% | 46952 : 80366<br>37% |
| 3a. Weight loss / decrease in appetite | AppDec | 9265 : 14594<br>39% | 39453 : 36497<br>52% | 0 : 0 |
| 3b. Weight gain / increase in appetite | AppInc | 7902 : 13167<br>38% | 22612 : 36489<br>38% | 0 : 0 |
| 4a. Insomnia | SleDec | 18917 : 6573<br>74% | 73144 : 19851<br>79% | 0 : 0 |
| 4b. Hypersomnia | SleInc | 10586 : 11050<br>49% | 20125 : 20055<br>50% | 0 : 0 |
| 5a. Psychomotor agitation | MotoInc | 10447 : 12372<br>46% | 113 : 3181<br>3% | 0 : 0 |
| 5b. Psychomotor slowing | MotoDec | 12701 : 11214<br>53% | 299 : 2995<br>9% | 0 : 0 |
| 6. Fatigue | Fatig | 23941 : 2497<br>91% | 85304 : 16736<br>84% | 0 : 0 |
| 7. Feelings of worthlessness / guilt | Guilt | 21921 : 3888<br>85% | 61757 : 43570<br>59% | 0 : 0 |
| 8. Diminished concentration | Conc | 23974 : 2386<br>91% | 75190 : 23416<br>76% | 0 : 0 |
| 9. Recurrent thoughts of death or suicide | Sui | 18170 : 9609<br>65% | 46984 : 58885<br>44% | 0 : 0 |

Data from the Psychiatric Genomics Consortium (PGC) was drawn from 23 cohorts in the Wave 1 and Wave 2 datasets of the Major Depressive Disorder Working Group (Major Depressive Disorder Working Group of the Psychiatric GWAS Consortium, 2013; Wray et al., 2018). Symptoms were assessed by trained interviewers using structured diagnostic instruments and DSM checklists. Because information on symptom presence was not available for control participants in most cohorts, participants with a diagnosis of depression were selected for analysis (N = 12,821).

The Australian Genetics of Depression Study (AGDS) (Byrne et al., 2020; Mitchell et al., 2022) is a study of depression and therapeutic response recruited using nationwide prescribing history and through publicity targeting adults who are or had ever been treated for clinical depression (N = 20,689). Symptoms experienced during the participant’s worst period of depression were assessed using the Composite International Diagnostic Interview (CIDI) Short Form (Hickie et al., 2001) and administered through an online questionnaire. Because the study was enriched for participants with a history of being diagnosed with or treated for depression, AGDS was grouped as a Clinical cohort.

Generation Scotland: Scottish Family Health Study (GS:SFHS) is a study of 7,000 families recruited from the general population of Scotland (Smith et al., 2012). Participants who screened reported seeking help for emotional or psychiatric problems were administered an in person structured interview (Fernandez-Pujals et al., 2015; Smith et al., 2012); and a subset participated in an online follow-up that included a CIDI (Composite International Diagnostic Interview) questionnaire. Symptom data was analysed on participants who met DSM criteria for depression at either time point (N = 3,493).

The Avon Longitudinal Study of Parents and Children (ALSPAC) is a UK-based population birth cohort (Boyd et al., 2013). Participants were from the children sample (N = 13,988) with symptoms present during the last two weeks assessed using the Clinical Interview Schedule Revised (CIS-R) (Lewis et al., 1992) collected during clinical visits at ages 18 and 24. Participants were considered to have had a symptom if they reported it at either measurement occasion.

Estonian Biobank (EstBB) is a population health cohort recruited from medical practitioners in Estonia (Leitsalu et al., 2015). Participants responded to a CIDI questionnaire of depression symptoms during the Mental Health online Survey (MHoS) recontact. Participants were first screened of the presence of low mood or anhedonia and then asked about symptoms during the worst period of depression (N = 84,079).

UK Biobank (UKB) is a population health cohort recruited from general practitioners in the United Kingdom (Sudlow et al., 2015). Lifetime depression symptoms were assessed during online recontact and taken from the CIDI portion of the Mental Health Questionnaire (Davis et al., 2020) (UKB-MHQ, N=157,366) and from assessments of low mood and anhedonia from the baseline touchscreen questionnaire (UKB Touchscreen, N=222,061). For the CIDI, low mood and anhedonia were used as gating symptoms, where participants had to endorse at least one to be asked about the other symptoms.

### Genome-wide association meta-analysis

Genome-wide association study (GWAS) analyses were conducted on each symptom separately in the cohorts (PGC, AGDS, GS:SFHS, ALSPAC, EstBB, UKB-MHQ) on participants who had genetic similarity with each other and the 1000 Genomes European reference. Participants in UKB who clustered with other reference populations were not analysed because sample sizes did not meet the threshold for LD score estimation (N > 5000). See Supplementary Material for more information on the individual study GWASs. We meta-analysed the GWAS summary statistics based on the ascertainment design. PGC, AGDS, and GS:SFHS were meta-analysed together to form the “Clinical” symptom summary statistics; and ALSPAC, EstBB, and UKB-MHQ were meta analysed together for the “Community” summary statistics. We performed the meta-analyses using Ricopili (Lam et al., 2020) and calculated SNP-based heritability using LD Score Regression (LDSC) (Bulik-Sullivan et al., 2015). For input into LDSC we set the sample size equal to the sum of effective sample sizes of each cohort in the meta-analysis and then specified sample prevalences of 50% (Grotzinger et al., 2022). Symptoms’ population prevalences were estimated for the Clinical cohorts by multiplying the observed sample prevalence by the prevalence of MDD (15%) and for the Community cohorts by multiplying by the proportion of participants in the UKB MHQ sample who were positive on either one the gating symptoms. We assessed significant associations in the meta-analysed summary statistics at *p* < 5 × 10^-8^ / 22 (the number of meta-analyses conducted) or at *p* < 5 × 10^-8^ with prior association or biological evidence at the locus.

### Confirmatory factor analysis of Genetic Covariance Structure

We fit confirmatory genetic factor analysis models to the meta-analysed ascertainment cohort (i.e., Clinical and Community-based) and UKB Touchscreen summary statistics for each symptom using Genomic SEM (Grotzinger et al., 2019). We first fit a common factor model, where all symptoms load on a single factor as a baseline, using symptoms with a non-negative LDSC heritability (Model A). To explore how sample ascertainment influenced the genetic correlations among the symptoms, we fit a series of models that captured various aspects of the sampling and measurement processes. We then used these results to inform the construction of models that grouped the symptoms based on previous findings and diagnostic criteria. We assessed relative model fit using Akaike Information Criterion (AIC) to pick the best model and absolute model fit with Standardized Root Mean Square Residual (SRMR) to determine how well the model was capturing the genetic correlations among symptoms. We also examined residual correlations to understand what aspects of symptom structure were not being captured. Factor structures are listed in Supplementary Table S4 and illustrated in Supplementary Figure S1.

### Ascertainment/measurement models

The most pertinent measurement difference among the symptoms was which meta-analysed cohorts the symptom came from, so we created a two-factor model where all symptoms from the same cohorts (Clinical or Community) loaded on the same factor (Model B). The next model considered the effect of the cardinal symptoms as gating items in UK Biobank and posited a general MDD factor that all the symptoms loaded on alongside an uncorrelated Gating factor with loadings from just the Community and UKB Touchscreen *low mood* and *anhedonia* symptoms (Model C). The Gating factor would therefore isolate variation associated with differences across the full non-clinical (dysphoria) to clinical spectrum. Symptoms not loading on the gating factor (i.e., those for which data are conditional on the presence of the two gating symptoms) represent variation within the more severe region of the spectrum and are thus more directly comparable to analyses of data from cases only. We then combined the Clinical Community and Gating models to create a three-factor model (Model D).

### Symptom models

Based off the best measurement model, we then fit models that grouped symptoms into two or three factors based on previous findings from phenotypic, twin, and Genomic SEM models and from diagnostic criteria. The two factor models grouped symptoms into Psychological and Somatic (Model E); Psychological and Neurovegetative (Model F); or Affective and Neurovegetative (Model G) factors (Elhai et al., 2012; Krause et al., 2008, 2010; Thorp et al., 2020). The Affective factor contained symptoms *low mood*, f*eelings of guilt,* and *suicidality*. The Psychological factors broadened the Affective factor to include the symptoms *anhedonia and/or loss of concentration*. The Somatic factor included the *appetite*, *sleep*, *fatigue,* and *psychomotor* symptoms. The Neurovegetative factors incorporated the somatic symptoms while also including *loss of concentration* and/or *anhedonia.* A three factor model (Model H) loaded symptoms onto cognitive (*feelings of guilt, loss of concentration, suicidality),* mood (*low mood, anhedonia, feelings of guilt),* and neurovegetative (*appetite, sleep, fatigue, psychomotor)* (Kendler et al., 2013).

We also fit factor models that disaggregated symptoms that involved an increasing or decreasing change (*appetite/weight, sleep, psychomotor*). One such model (Model I) was based on previous findings that identified factors for Appetite (*appetite/weight decrease* and *increase),* vegetative (*hypersomnia, psychomotor slowing, fatigue, concentration)* and Cognitive/Mood (*low mood, anhedonia, insomnia, psychomotor agitation, feelings of guilt, suicidality)* (van Loo et al., 2022). Finally, we considered a three-factor model (Model J) based on diagnostic criteria of melancholic depression (*anhedonia, insomnia, psychomotor agitation, appetite/weight decrease, feelings of guilt*) and atypical depression (*hypersomnia, appetite/weight increase, psychomotor slowing, fatigue*), with the remaining symptoms loading on an Affective/Cognitive factor (*low mood, suicidality, loss of concentration)*.

### Genomic factor meta-analysis

We conducted multivariate meta-analyses of symptoms in Genomic SEM (Grotzinger et al., 2019). Because of low power in some of the Clinical and Community symptoms summary statistics, we were unable to fully test SNP effects on symptom factors. We therefore fit a model with single common factor meta-analysis across well-powered symptoms 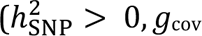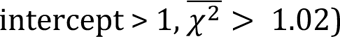 and tested for SNP heterogeneity at the level of individual symptoms. A genomic factor meta-analysis estimates SNP associations in two structural models: a common pathway model where the SNP is associated with each symptom through its effect on the factor and an independent pathway model where the SNP is associated independently with each symptom, bypassing the common factor. The SNP coefficients from the common pathway model act as a meta-analysis of the symptom summary statistics while accounting for sample overlap. A comparison of fit between the common and independent pathway models yields a heterogeneity statistic for each SNP, QSNP, indicating whether the SNP’s association varies between symptoms.

### Regression Analysis

Using the best fitting models, we tested how the factors were related to correlates of depression. We selected phenotypes that are known to genetically correlate with depression, including psychiatric disorders (anxiety disorder, bipolar disorder, PTSD, schizophrenia); depression defined through clinical ascertainment (major depressive disorder) and through broader or more minimal definitions (major depression); and other health, behavioural, and social phenotypes (see Supplementary Materials for list of studies). We tested whether the other phenotypes had specific genetic correlations with each symptom factor. We did this by first fitting single regressions of a phenotype on each symptom factor. We then compared this to a multiple regression of the phenotype on all symptom factors simultaneously. We used Benjamini–Yekutieli FDR adjustment to correct for multiple testing (Benjamini & Yekutieli, 2001).

## Results

### Genome-wide association and meta-analyses

We conducted GWAS for each symptom separately in all cohorts and meta-analysed within sample ascertainment groups (*Clinical* or enriched cohorts: PGC, AGDS, GS:SFHS; *Community* cohorts: ALSPAC, EstBB, UKB-MHQ) (Supplementary Table S1). Two associations met the stringent multiple testing burden (*p* < 5 × 10^-8^ / 22). One was an intron in *FTO* (ENSG00000140718, alpha-ketoglutarate dependent dioxygenase, a gene involved in food intake) associated with *Weight gain* in the Community cohorts. The other was associated with *Anhedonia* in the Community cohorts and was an intron variant in an uncharacterised non coding RNA gene (LOC105379109/ENSG00000251574) and in a region previously associated with neuroticism, depression, and subjective well-being.

At the genome-wide significance threshold (*p* < 5 × 10^-8^) there were three associations that were also supported by prior evidence. There were two associations with *Depressed mood* in the Community cohorts: an intron in COMP (ENSG00000105664, cartilage oligomeric matrix protein) also near *CRTC1* (ENSG00000105662, CREB regulated transcription coactivator 1, a gene that regulates metabolism); and an intron in an uncharacterised gene (LOC107986777) regionally associated with depression. An upstream variant for an uncharacterised long intergenic non-protein coding RNA (LINC01938) was associated with Community *Anhedonia* and in a region previously associated with neuroticism and major depressive disorder.

LDSC-estimated heritabilities were primarily in the 0.025–0.1 range (Figure 1, Supplementary Table S3). Many of the symptoms in the Clinical cohorts (*Depressed mood, Anhedonia, Fatigue, Concentration)* had negative heritabilities and the psychomotor symptoms from the Community cohorts did not meet the sample size inclusion criteria (NEff > 5000).

**Figure 1.**
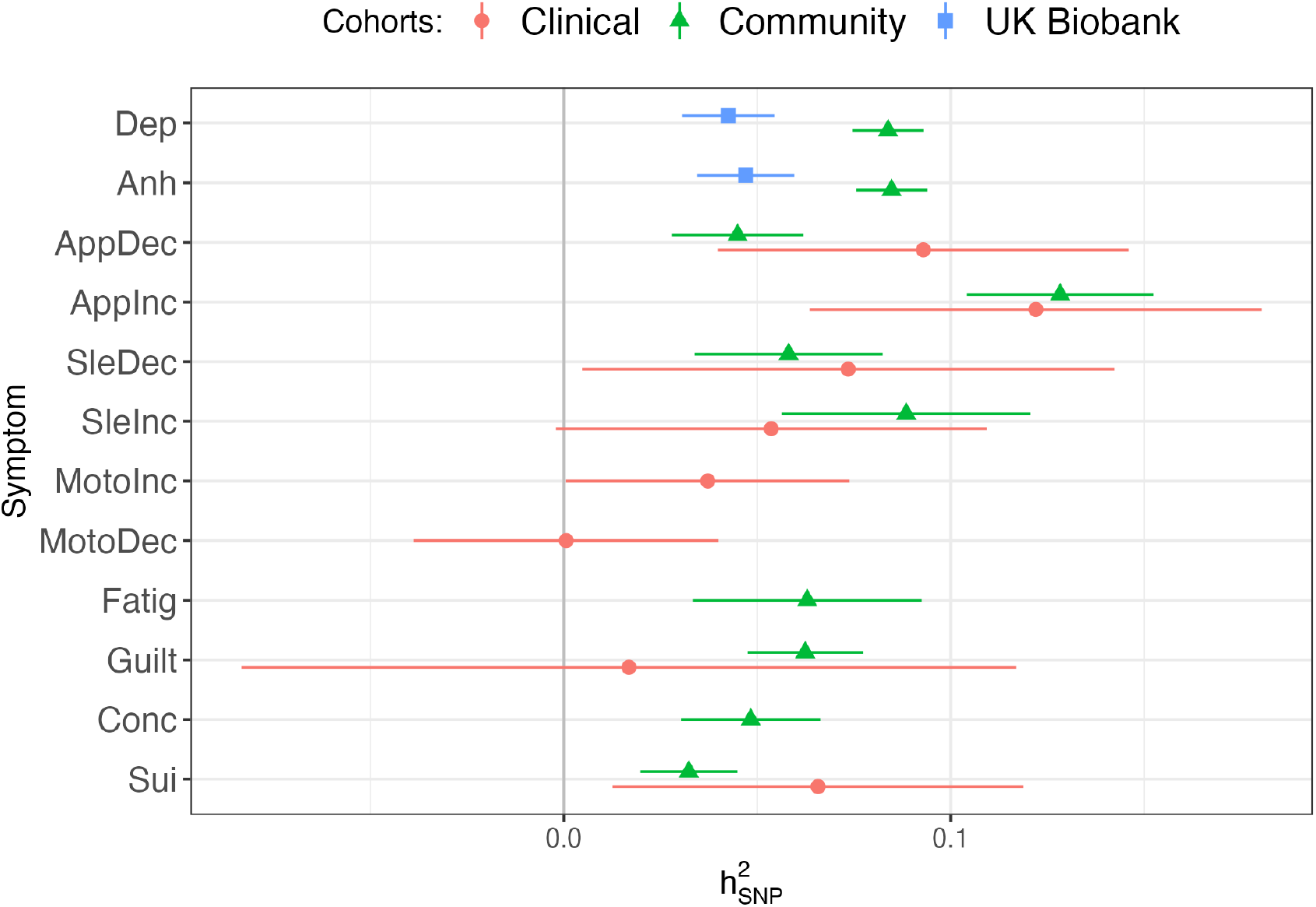
LDSC-estimated heritabilities 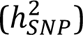 on the liability scale of depression symptoms (abbreviations are listed in Table 1) for summary statistics that met inclusion criteria 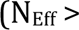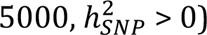 Clinical = PGC + AGDS + GS:SFHS meta-analysis, Community = ALSPAC + EstBB + UKB-MHQ meta-analysis, UK Biobank = UKB-Touchscreen GWAS.

### Confirmatory factor analysis

We brought forward symptoms from the Clinical and Community cohorts meta-analyses and the UKB Touchscreen assessment that had a 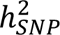 greater than 0 and sample sizes > 5000 for confirmatory factor analysis (Supplementary Table S4, Supplementary Figure S1a–l).

A common factor model (A) of the symptoms showed poor fit (CI=0.932, SMR=0.169, AIC = 5355). A model (B) with separate factors for Clinical and Community cohort symptoms had slightly poorer fit (AIC = 5369) and yielded a genetic correlation between the two factors of *rg* = 0.63±0.14, p = 1.3 ×10^-5^. An alternative model (C) that only split off the Community and UKB Touchscreen *mood* and *anhedonia* symptoms into an orthogonal factor, capturing these symptoms use as gating items in EstBB and UKB-MHQ, showed substantially improved fit (AIC = 3229). A model (D) combining the sample factors with the orthogonal Gating factor also improved model fit (AIC = 3285) and led to a nominal increase in the genetic correlation between the Clinical and Community factors to *rg* = 0.75±0.17, p = 6.9 ×10^-6^.

We then tested whether models that grouped symptoms together across cohorts fit better than the factor models based on sampling methodology. Because the addition of an orthogonal Gating factor improved model fit so substantially, the symptom-oriented models all included this factor. The best fitting of the symptom models was Model I which included factors capturing Appetite, Vegetative, and Cognitive/Mood symptoms (Figure 2). The Appetite factor had a genetic correlation of *rg* = 0.66±0.09 (p = 1.4 ×10^-12^) with the Vegetative factor and *rg* = 0.48±0.07 (p = 3.4 × 10^-13^) with the Cognitive/Mood factor, while the Vegetative and Cognitive/Mood factors were more highly correlated, *rg* = 0.91±0.07 (p = 7.8 × 10^-40^).

**Figure 2.**
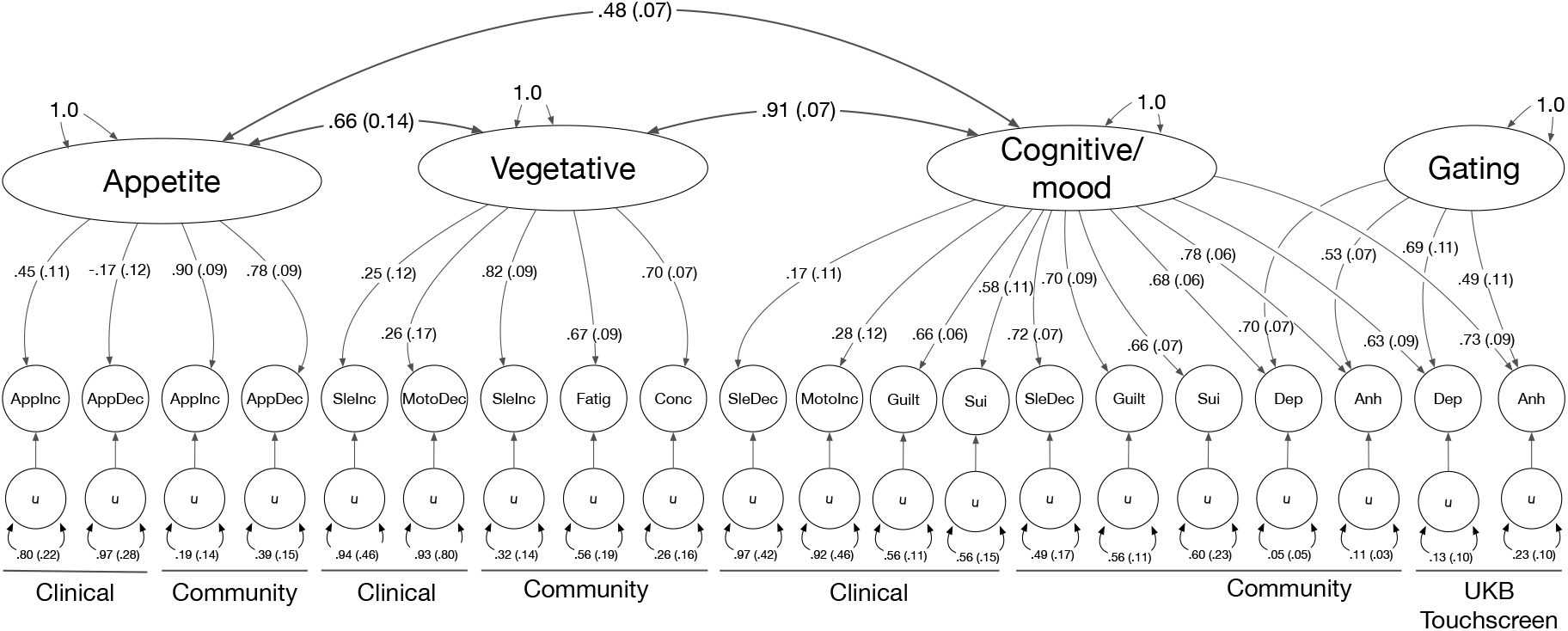
Standardised loadings (standard errors) of factors on symptoms and genetic correlations among factors for the best fitting model (Model I). Symptom abbreviations are listed in Table 1.

None of the models fully captured the genetic correlations among the symptoms, as indicated by high SRMR (Model D = 0.149, Model I = 0.147). An inspection of the residual genetic correlations (Supplementary Figure S3d) indicated correlations between the same symptoms across the two cohorts (e.g., Clinical *appetite decrease* with Community *appetite decrease*) were not fully represented by the factor structure. We thus tested how adding residual correlations between symptoms that included from both cohorts (*appetite decrease, appetite increase, insomnia, hypersomnia,* and *suicidality)* improved absolute model fit. The addition of these residual correlations lowered SRMR to 0.140.

### Multivariate meta-analysis of symptoms

Because many of the symptom summary statistics were low powered, we were unable to conduct a multivariate meta-analysis using the genetic factors. Alternatively, to test for SNP effects specific to each symptom, we conducted a multivariate meta-analysis of well-powered symptoms summary statistics. The common factor had three genome-wide significant associated variants: an intron variant in BRINP2 (ENSG00000198797, BMP/retinoic acid inducible neural specific 2, a regulator of neuronal differentiation); the upstream variant that was associated with Community *Anhedonia* symptoms; and an intron variant in LRRC37A3 (ENSG00000176809, leucine rich repeat containing 37 member A3) An intron in the FTO gene showed substantial heterogeneity in the common factor meta-analysis (Supplementary Table S7).

### Genetic multiple regression

To determine whether each MDD symptom factor had specific genetic relationships with twelve phenotypes, we conducted genetic multiple regressions in GenomicSEM using the Clinical– Community sample and Appetite–Vegetative–Cognitive/Mood symptom factor models. Because the Vegetative and Cognitive/Mood symptom factors had a high genetic correlation, we combined these into one factor, which we labelled Depression. We first calculated single regressions of each phenotype on each of the factors separately, where the standardized coefficient indicated the overall shared genetics with each factor. We then then fit a genetic multiple regression with the two models, where the standardized coefficient represents the unique genetic relationship of the phenotype with each factor, after adjusting for shared overlap with the other factors (e.g.., a phenotype’s genetic relationship with the Clinical factor after adjusting for the Community factor, and vice versa; and its genetic relationship with the Appetite factor after adjusting for the Depression and Gating factors, and vice versa). In the single regression (unadjusted) analysis, the genetic relationship of each phenotype with all of the factors were in the same direction with the exception of educational attainment which had a negative relationship with most of the factors (at *p* < 0.0005) but a positive yet non-significant relationship with the Gating factor (Figure 3, Supplementary Table S7). After adjusting for shared overlap with the Clinical factor, the Community factor did not have any specific relationships with the phenotypes, while the Clinical factor had specific, positive genetic correlations with anxiety, bipolar disorder, major depression, major depressive disorder, neuroticism, PTSD, and chronic pain.

**Figure 3.**
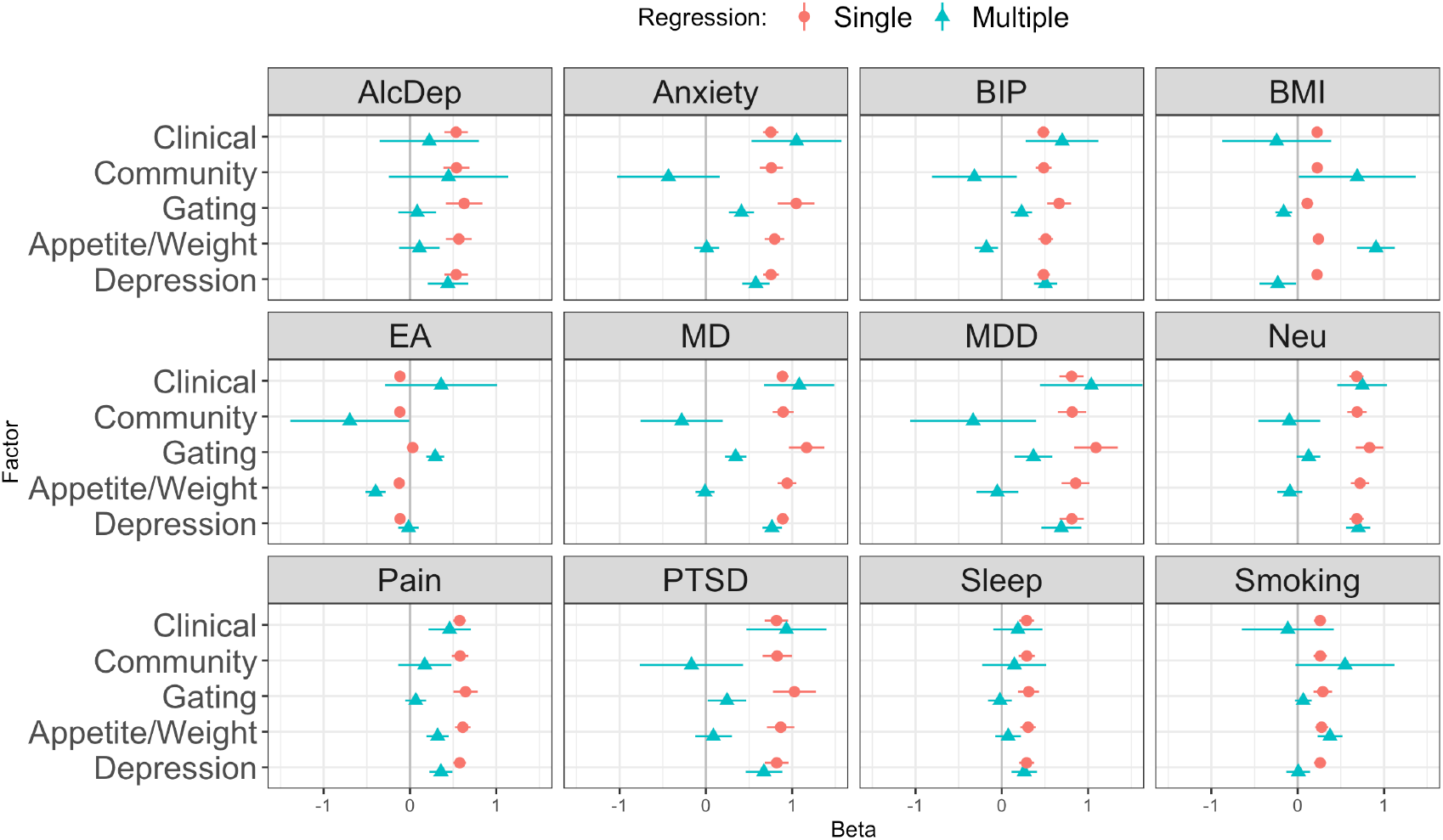
Genetic regression of phenotypes on Clinical and Community cohort factors; and on Gating, Appetite/Weight, and Depression symptom factors. Single genetic regression standardized beta coefficients (green triangles) and multiple genetic regression coefficients, which adjust for the other factors (Clinical-Community adjusted together; Gating-Appetite-Depression adjusted together). AlcDep = alcohol dependence, Anxiety = anxiety disorder, BIP = bipolar disorder, BMI = body-mass index, EA = educational attainment, MD = major depression, MDD = major depressive disorder, Neu = neuroticism, Pain = chronic pain, PTSD = post-traumatic stress disorder, Sleep = long sleep duration, Smoking = cigarettes per day.

When adjusting for the Depression and Gating factors, the factor for the Appetite symptoms had specific genetic correlations with BMI and smoking. The factor for the remaining Depression symptoms was specifically correlated with alcohol dependence, anxiety, neuroticism, and long sleep duration. The Gating factor did not have any specific genetic correlates after adjusting for the other factors, but it did share with the Depression factor positive genetic correlations with bipolar disorder, major depression and major depressive disorder, and PTSD after adjustment for the Appetite factor (Figure 3, Supplementary Table S7).

## Discussion

We used genome-wide association data to analyse the genetic relationships among symptoms of depression based on cohort sampling and symptom content and to estimate whether the genetic factors had specific correlates with other phenotypes. We analysed data from two sets of cohorts: Clinical cohorts that were ascertained to have depression through clinical or interview assessments or were recruited preferentially on a history of treatment for depression; and Community cohorts that were not recruited based on disease status (but for which symptom data was typically conditioned based on endorsement of cardinal gating symptoms). We conducted GWAS of major depression symptoms in each cohort then meta-analysed within the Clinical and Community groups.

We identified loci associated with individual major depression symptoms and with a common genetic factor of the symptoms. Several associations from the individual symptoms and common factor meta-analysis (rs7515828, r rs30266, s6884321, rs150046352) have been identified previously in GWAS of or unipolar depression (EFO ID EFO_0003761) (Sollis et al., 2023) or in meta-analyses of major depressive disorder (Als et al., 2022; Howard et al., 2019; Levey et al., 2021; Wray et al., 2018) SNPs associated with *Appetite / weight increase* have primarily come up in GWAS of body mass index and related traits (Elsworth et al., 2020; Hoffmann et al., 2018; Howe et al., 2022; Yengo et al., 2018) but another SNP in the FTO gene has also been associated with atypical subtypes (Milaneschi et al., 2014).

Many symptoms in the Clinical cohorts had heritability estimates that were zero or negative. This is not unexpected. Selecting individuals based on their phenotype (that is, that they are or have been affected by depression) would result in a sample that has a lower genetic variance for traits that contribute to the determination of the phenotype (Falconer & Mackay, 1996). Additionally, the symptoms with negative heritabilities (*depressed mood, anhedonia, fatigue, concentration problems* in the clinical cohorts) also had high endorsement rates (85–94%), and power to detect heritability is reduced the further the sample prevalence deviates from 50% (Lee et al., 2011). In contrast, the other symptoms had more equal endorsement rates (34–75%) among affected participants. Although the low heritabilities of symptoms from the Clinical cohorts limited the comprehensiveness of alternative factor models that could be tested, we did find congruence between the Clinical and Community cohort symptoms, with a high genetic correlation between their respective factors. We also showed that model fit was substantially improved by modeling the use of cardinal symptoms (*Low mood* and *Anhedonia*) as gating items for surveys of depression symptoms. Among the models that grouped symptoms together without consideration for symptom direction, we found broad support for a split between psychological and somatic symptoms identified in previous phenotypic (Elhai et al., 2012) and genetic (Thorp et al., 2020) analyses. When directional symptoms were portioned out based on diagnostic specifiers, we found that a three-factor model capturing Appetite, Vegetative, and Cognitive/Mood symptoms (van Loo et al., 2022) had the best fit among all models considered. The correlations among the factors indicated that the Vegetative and Cognitive/Mood symptoms should be grouped together, with only the Appetite symptoms making up a possibly different dimension of depression. However, the Clinical cohort symptoms had low loadings in both the sample-based and symptom-based models (except for the Clinical *Appetite/Weight* and *Suicidality* symptoms), and thus the model fit was driven primarily by capturing the structure among the Community cohort symptoms. This observation is consistent with the fact that the Clinical cohorts are more selected than the community cohorts, and that conditioning data presence on a diagnosis can induce downward bias in correlations amongst the symptoms that aggregate to form the diagnosis. Similar attenuation, albeit to a lesser degree, may be expected for items in community samples whose presence was conditioned on endorsement of cardinal symptoms.

Despite these limitations, the Clinical factor was genetically correlated with all the other phenotypes selected for comparison. A multiple genetic correlation analysis showed that the Clinical and Community factors had a shared genetic relationship with alcohol dependence, bipolar disorder, BMI, educational attainment, MDD, chronic pain, sleep, and smoking. The symptom-based factor model showed discriminative validity between the Appetite symptoms and the rest of the symptoms of depression through relationships with correlates of depression. A positive genetic correlation between increase in appetite/weight with BMI has previously been shown with PGC cohorts (Milaneschi et al., 2017) and in UKB (Badini et al., 2022), and our findings show that this result holds even when adjusting for genetic overlap with other symptoms.

Our results demonstrate the challenges and insights associated with considering symptoms of depression separately. In particular, substantial care must be taken to consider how samples are ascertained (clinical versus community recruitment), how symptoms are measured (the use of gating items in symptom inventories), and including assessments of item direction (e.g., insomnia versus hypersomnia) when modelling the genetic structure of depression symptoms. However, the evaluation of direction was limited to a small subset of symptoms and did not include distinctions such, as low versus irritable mood, or included only partial assessments, such as weight but not appetite changes being assessed in UKB. The coverage of features of atypical and melancholic depression was likewise incomplete. For example, several diagnostic features of the atypical specifier were not included, such as mood reactivity, sensitivity to interpersonal rejection, and leaden paralysis. We also only examined subtypes defined by symptom profiles and not other sources of heterogeneity such as onset, life event exposure, or treatment outcomes (Harald & Gordon, 2012) which may also have a differential biological and genetic basis (Beijers et al., 2019; Milaneschi et al., 2020; Nguyen et al., 2022). The strongest genetic associations were between symptoms of weight/appetite change and genes linked to satiety and metabolism. This highlights the need to phenotype somatic symptoms (weight or sleep changes and fatigue) outside of the context of mental health assessments, so that their specific role in depression can be better isolated. Likewise, the use of gating symptoms makes it difficult to fully capture the range of genetic risk between everyday dysphoria and differences among affected individuals. While the results support the idea that depression is heterogeneous, the genetic liability for symptom profiles and comorbidities can be captured in relatively few dimensions.

## Data Availability

Meta-analysed summary statistics are available for download from the PGC website. Individual-level PGC data is available by application to the PGC Data Access Committee. Data from Estonian Biobank, UK Biobank, and ALSPAC are available to bonafide researchers upon application. Data from AGDS is available for collaboration by contacting NGM.

https://www.med.unc.edu/pgc/download-results

https://www.med.unc.edu/pgc/shared-methods/

https://genomics.ut.ee/en/content/estonian-biobank

https://www.ukbiobank.ac.uk

http://www.bristol.ac.uk/alspac/

## Code and data availability

Primary code is available from the Psychiatric Genomics Consortium (PGC) GitHub Repository (https://github.com/psychiatric-genomics-consortium/mdd-symptom-gwas/) and meta analysed summary statistics are available for download from the PGC website (https://www.med.unc.edu/pgc/download-results/). Individual-level PGC data is available by application to the PGC Data Access Committee (https://www.med.unc.edu/pgc/shared-methods/). Data from Estonian Biobank (https://genomics.ut.ee/en/content/estonian-biobank), UK Biobank (https://www.ukbiobank.ac.uk) and ALSPAC (http://www.bristol.ac.uk/alspac/) are available to bonafide researchers upon application. Data from AGDS is available for collaboration by contacting NGM.

## Acknowledgments

AGDS supported by National Health and Medical Research Council (NHMRC) (1086683, 1145645, 1078901, 1087889, 1173790); ALSPAC supported by the Medical Research Council and Wellcome Trust (217065/Z/19/Z). Estonian Biobank supported by the European Union through the European Regional Development Fund (Project No. 2014-2020.4.01.15-0012). NTR-NESDA supported by Biobanking and Biomolecular Resources Research Infrastructure (BBMRI-NL; 184.021.007 and 184.033.111), National Institutes of Health (NIH, R01D0042157-01A, MH081802, Grand Opportunity grants 1RC2 MH089951 and 1RC2 MH089995). Part of the genotyping and analyses were funded by the Genetic Association Information Network (GAIN) of the Foundation for the National Institutes of Health. Funding for NTR is acknowledged from NWO-MW 904-61-193; NWO 985-10-002; NWO 904-61-090; Royal Netherlands Academy of Science Professor Award (PAH/6635) to DIB; European Research Council (ERC-230374).

Funding for the infrastructure of the NESDA study (www.nesda.n) was obtained from the Netherlands Organization for Scientific Research (Geestkracht program grant 10-000-1002); the Center for Medical Systems Biology (CSMB, NWO Genomics), VU University Medical Center, GGZ inGeest, Leiden University Medical Center, Leiden University, GGZ Rivierduinen, University Medical Center Groningen, University of Groningen, Lentis, GGZ Friesland, GGZ Drenthe, Rob Giel Onderzoekscentrum. MJA, ASFK, and AMMc are supported by the Wellcome Trust (104036/Z/14/Z, 220857/Z/20/Z). SEM is supported by NHMRC APP1172917, APP1138514 and MRF1200644. ADG, and MGN and EMTD were supported by National Institute of Mental

Health grant R01MH120219. KL and KK were supported by the Estonian Research Council grant PSG615. UKB analysis conducted under project 4844. This work made use of the NL Genetic Cluster Computer (http://www.geneticcluster.org) hosted by SURFsara and resources provided by the Edinburgh Compute and Data Facility (ECDF) (http://www.ecdf.ed.ac.uk/). The Psychiatric Genomics Consortium (PGC) has received major funding from the National Institute of Mental Health and the National Institute on Drug Abuse (U01 MH109528, U01 MH109532, U01 MH094421, U01 MH085520). This paper represents independent research part-funded by the NIHR Maudsley Biomedical Research Centre at South London and Maudsley NHS Foundation Trust and King’s College London.

For the purposes of open access, the author has applied a Creative Commons Attribution 4.0 International Public License (CC BY 4.0) to any Accepted Author Manuscript version arising from this submission.

## Declarations

CL sits on the SAB for Myriad Neuroscience, has received consultancy fees from UCB, and speaker fees from SYNLAB. HJG has received travel grants and speaker’s honoraria from Fresenius Medical Care, Neuraxpharm, Servier and Janssen Cilag as well as research funding from Fresenius Medical Care.

## Supplementary text: Genetic structure of major depression symptoms in clinical and population cohorts

### Genotyping and QC

#### Psychiatric Genomics Consortium (PGC)

The analysis used data from 24 cohorts from the PGC MDD datasets that had symptom data on cases. Data was drawn from the following cohorts:

- BiDirect (bidi1)
- BOMA (boma)
- CoFams (cof3)
- PsyCoLaus (col3)
- GenRED (gens, grnd)
- GenPod/Newmeds (gep3)
- GSK (gsk2)
- Jannsen (janpy)
- MPIP/MARS (mmi2, mmo4)
- NESDA/NTR (nes1)
- QIMR (qi3c, qi6c, qio2)
- RADIANT (rad3, rage, rai2, rau2, rde4)
- Rotterdam (rot4)
- SHIP (shp0)
- STAR*D (stm2)
- TwinGene (twg2)

The genotypes were processed through Ricopili (Lam et al., 2020) with the following QC: SNP missingness < 0.05; sample missingness < 0.02; autosomal heterozygosity deviation (|Fhet|<0.2); and SNP Hardy-Weinberg equilibrium (P>10^−6^ in controls, P>10^−10^ in cases). QC’d genotypes were then imputed to the 1000 Genomes Reference Panel (The 1000 Genomes Project Consortium, 2015). Information on cohort genotyping and additional processing steps is available in (Wray et al., 2018).

#### Australian Genetics of Depression Study (AGDS)

Genotyping was conducted using the Illumina Infinium Global Screening Array platform and QC’d for unknown or ambiguous map position and strand alignment, missingness >5%, HWE < 1 ×10^-6^, MAF<1%. Genotypes were imputed to HRCr1.1. Individuals were excluded with missing rate > 3%, inconsistent sex, or if deemed ancestry outliers from the European population (6 standard deviations from the first two genetic principal components from 1000 Genomes). Imputed genotype dosages were used for the analyses. GWAS was carried out in SAIGE (Zhou et al., 2018) using a generalized linear mixed model with genotyping batch and 10 PCs as covariates. Variants with MAF<1% and imputation accuracy score <0.7 were excluded.

#### Avon Longitudinal Study of Parents and Children (ALSPAC)

ALSPAC children were genotyped using the Illumina HumanHap550 quad chip genotyping platforms. Individuals were excluded on the basis of gender mismatches; minimal or excessive heterozygosity; disproportionate levels of individual missingness (>3%) and insufficient sample replication (IBD < 0.8). Population stratification was assessed by multidimensional scaling analysis, removing samples that clustered outside the CEU HapMap2 population. SNPs with a minor allele frequency of < 1%, a call rate of < 95% or evidence for violations of Hardy-Weinberg equilibrium (P < 5E-7) were removed. Cryptic relatedness was measured as proportion of identity by descent (IBD > 0.1). Related subjects that passed all other quality control thresholds were retained during subsequent phasing and imputation. 9,115 subjects and 500,527 SNPs passed these quality control filters. Imputation of the target data was performed using Impute V2.2.2 against 1000 genomes reference panel (Phase 1, Version 3) (all polymorphic SNPs excluding singletons), using all 2186 reference haplotypes (including non-Europeans). This resulted in 28,699,419 SNPs, with 8,282,911 SNPs with a MAF >0.01 and info score of >0.8. Analysis were conducted using SNPTEST v2.5.2, adjusting for sex and the first 10 principal components of ancestry.

#### Generation Scotland (GS:SFHS)

GWAS data was obtained using the Illumina OmniExpress array, and imputed using the Haplotype Research Consortium (HRC) dataset. Further details of methods here https://pubmed.ncbi.nlm.nih.gov/28270201/. GWAS was conducted in regenie with 4 PCs removing SNPs with MAC < 100, genotype missingness > 10%, INFO < 0.1, and HWE p > 1e-15.

#### Estonian Biobank (EstBB)

The samples from the Estonian Biobank have been genotyped at the Genotyping Core Facility of the Institute of Genomics, University of Tartu using the Global Screening Array (GSAv1.0, GSAv2.0, and GSAv2.0_EST) from Illumina. Altogether 155,772 samples have been genotyped and PLINK format files exported using GenomeStudio v2.0.4. Individuals were excluded from the analysis if their call-rate was <95% or if the sex defined based on heterozygosity of the X chromosome did not match the sex in the phenotype data. Variants were excluded if the call-rate was < 95% and HWE p-value <1e-4 (autosomal variants only). Variant positions were updated to genome build 37 and all alleles were switched to the TOP strand using tools and reference files provided at https://www.well.ox.ac.uk/~wrayner/strand/. After QC the dataset contained 154,201 samples for imputation. Before imputation variants with MAF<1% and indels were removed. Prephasing was done using the Eagle v2.3 software. The number of conditioning haplotypes Eagle2 uses when phasing each sample was set to: --Kpbwt=20000. Imputation was done using Beagle v.28Sep18.793 with effective population size ne=20,000. An Estonian population specific imputation reference of 2,297 WGS samples was used. The analysis was performed using the SAIGE software, including related individuals and adjusting for the first 10 principal components (PCs) of the genotype matrix, as well as for birth year, birth year squared and sex.

#### UK Biobank (UKB)

Imputed genotypes were analysed from the version 3 release (Bycroft et al., 2018). Imputed genotypes were QC’d to INFO >= 0.1, MAC >= 100, HWE P > 1e-10, max alleles = 2, and duplicate markers removed. Association analysis was performed as a logistic regression in Plink2 (Chang et al., 2015) with genotyping array and 20 PCs as covariates.

### Ethics statements

Ethical approval was obtained from the ALSPAC Ethics and Law Committee and the local research ethics committees (project number B3118). Consent for biological samples has been collected in accordance with the Human Tissue Act (2004). GWAS data was generated by Sample Logistics and Genotyping Facilities at Wellcome Sanger Institute and LabCorp (Laboratory Corporation of America) using support from 23andMe.

All participants in AGDS provided informed consent that they had read and understood the study information sheets and to confirm that they would be willing to provide a saliva sample for genotyping and downstream generic analyses. All study protocols were approved by the QIMR Berghofer Medical Research Institute Human Research Ethics Committee - approval numbers P2118, P1309 and P2304.

The activities of the EstBB are regulated by the Human Genes Research Act, which was adopted in 2000 specifically for the operations of the EstBB. Individual-level data analysis in the EstBB was carried out under ethical approvals [1.1-12/2860 & 1.1-12/624] from the Estonian Committee on Bioethics and Human Research (Estonian Ministry of Social Affairs), using data according to the release application [3-10/GI-28207] from the Estonian Biobank.

Ethical approval for the GS:SFHS data collection was obtained from the Tayside Committee on Medical Research Ethics A (ref 05/S1401/89). Generation Scotland is currently approved as a Research Tissue Bank by the East of Scotland Research Ethics Service (ref 20/ES/0021).

UK Biobank received ethical approval from the Research Ethics Committee (reference 11/NW/0382).

### Confirmatory factor analysis model schematics

Supplementary Figure S1. Schematic drawings of the CFA models

Schematics to illustrate factor structures of the models that were tested. See main text Table 1 for symptom abbreviations and Supplementary Tables S4,6 for factor structures and coefficients.

**Figure S1a:**
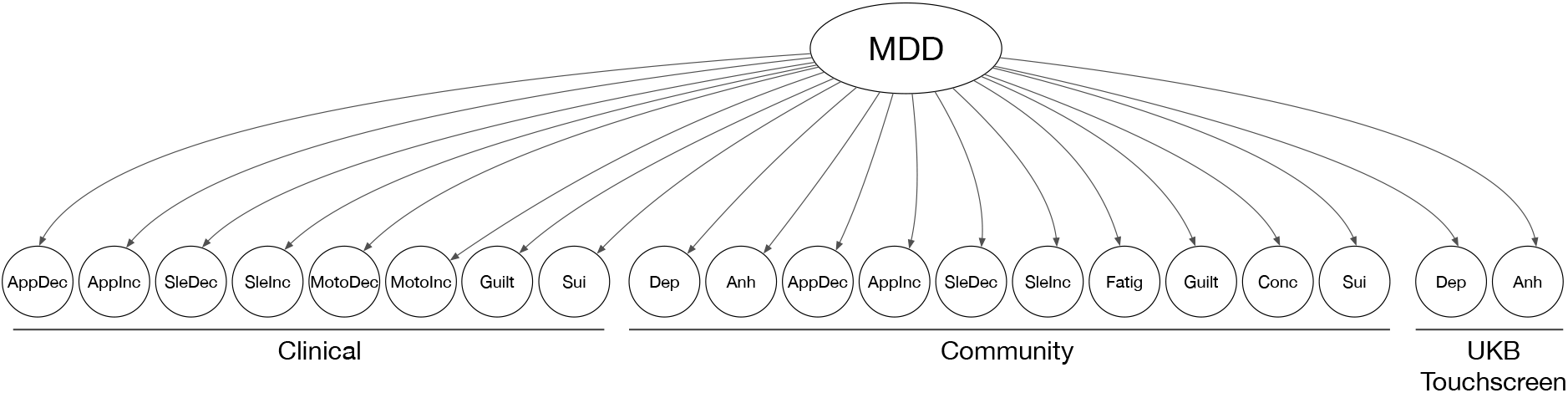
Model A: Common factor

**Figure S1b:**
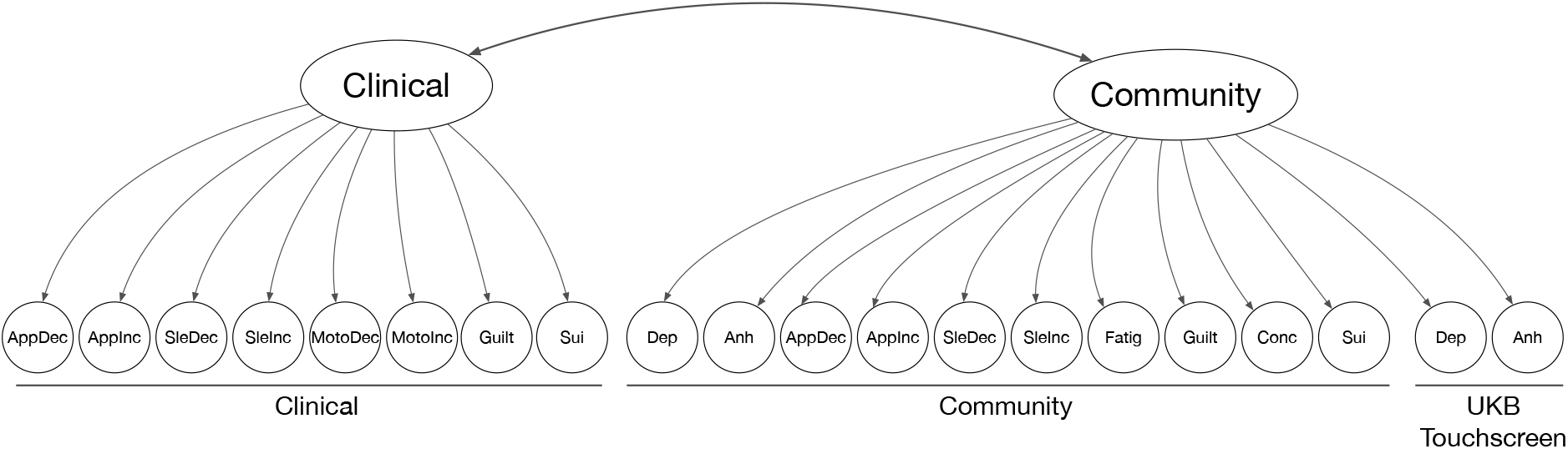
Model B: Clinical and community factors

**Figure S1c:**
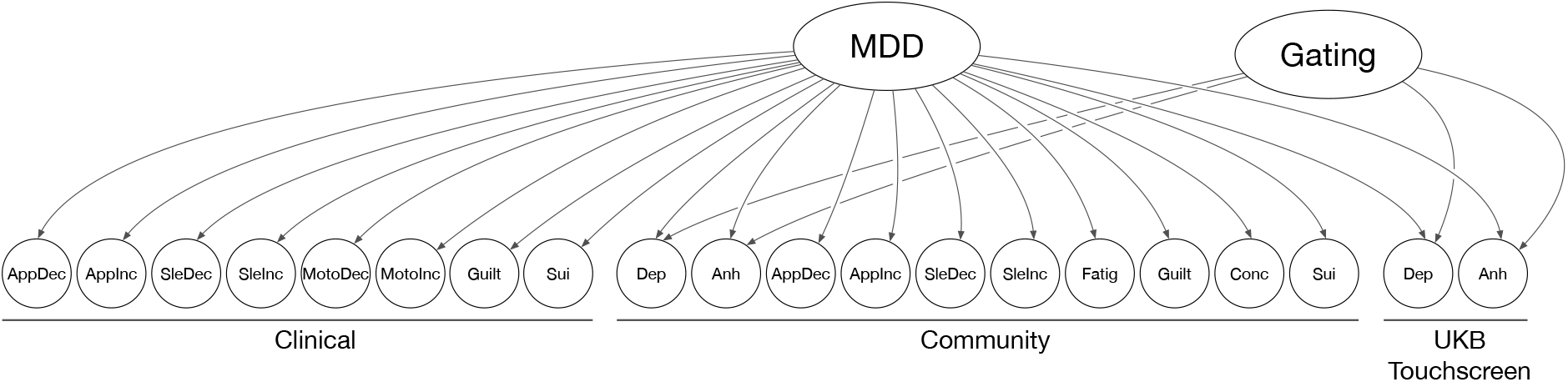
Model C: Gating measurement factor

**Figure S1d:**
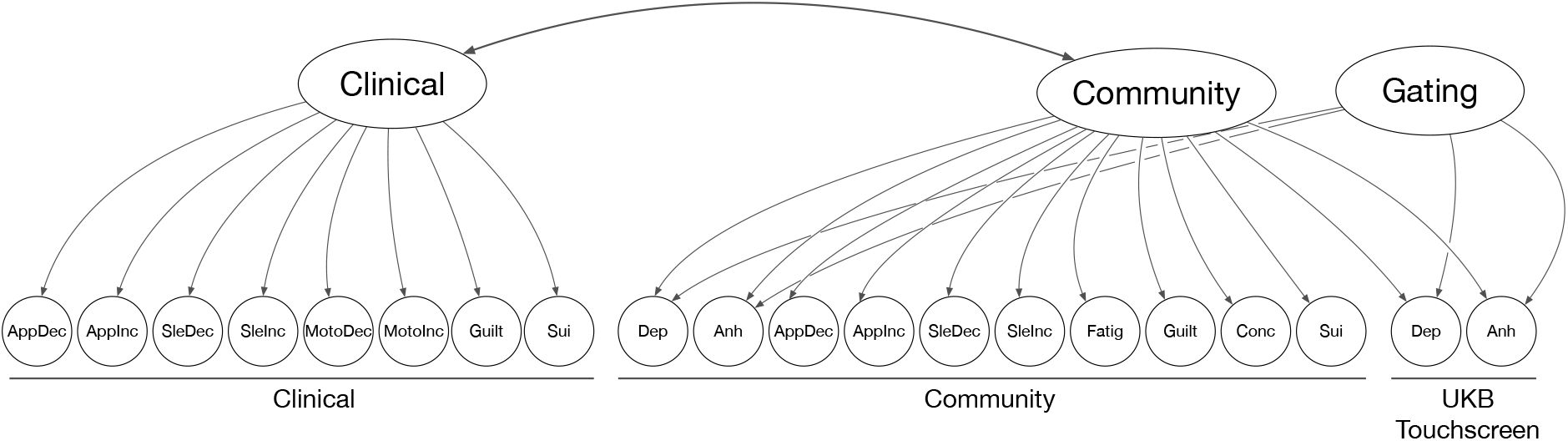
Model D: Clinical-Community-Gating factors

**Figure S1e:**
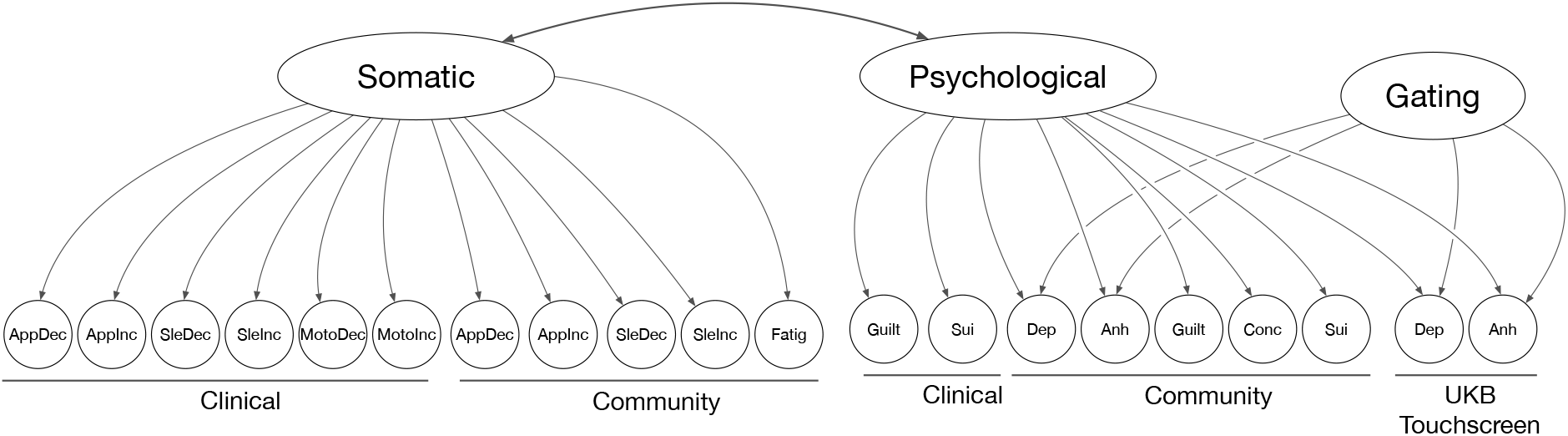
Model E: Psychological-Somatic

**Figure S1f:**
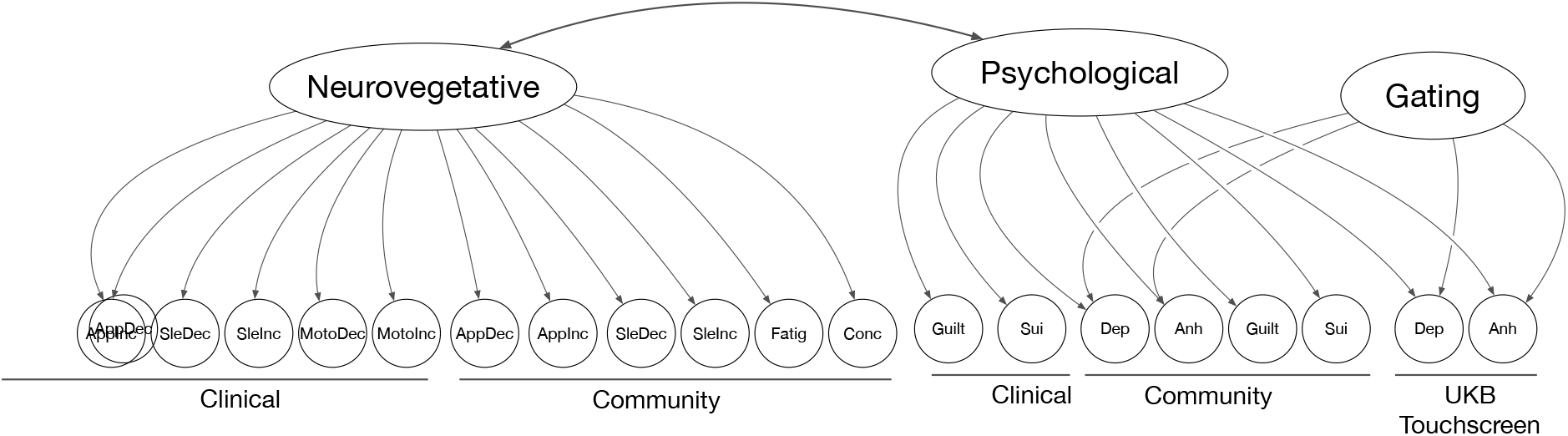
Model F: Psychologicals Neurovegetative

**Figure S1g:**
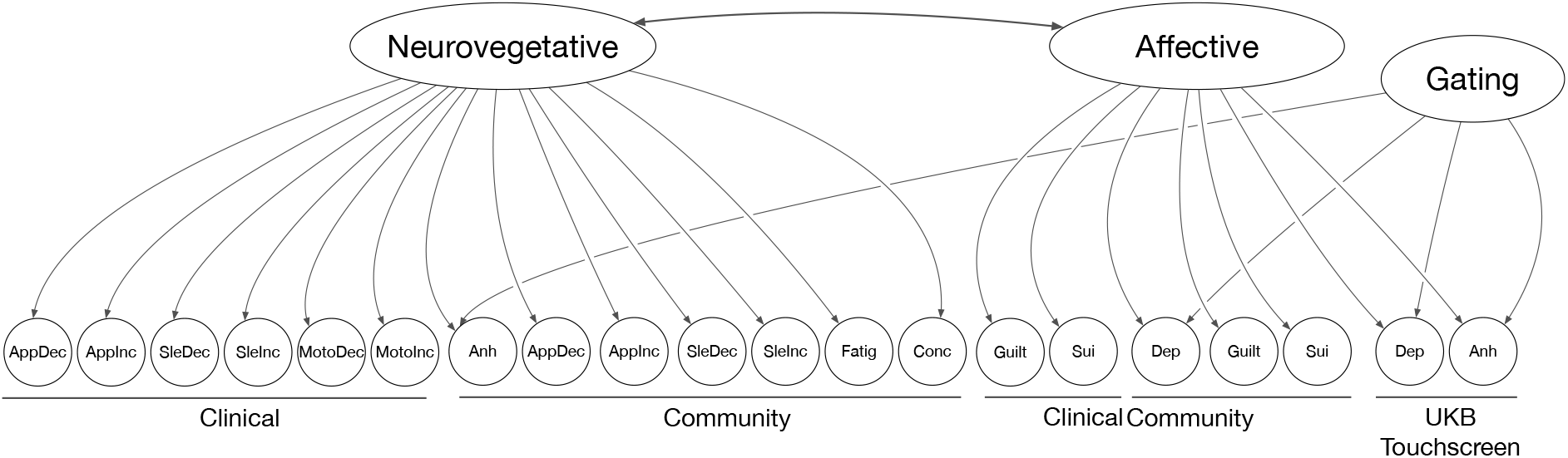
Model G: Affective-Neurovegetative

**Figure S1h:**
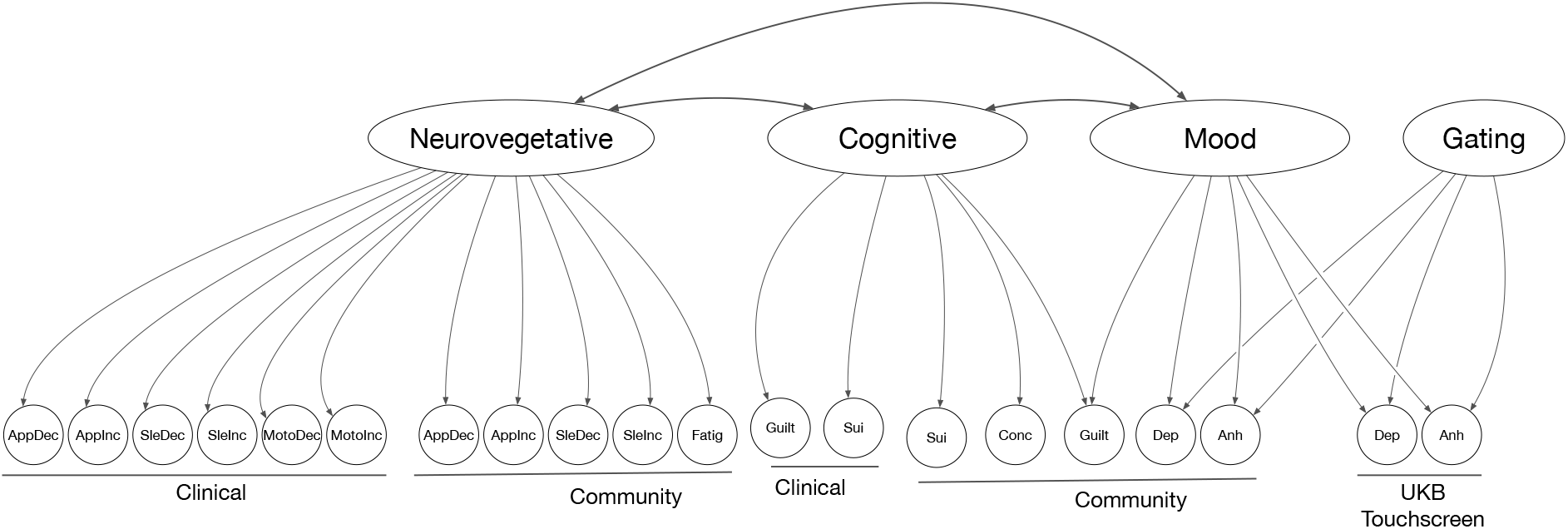
Model H: Cognitive-Mood-Neurovegetative

**Figure S1i:**
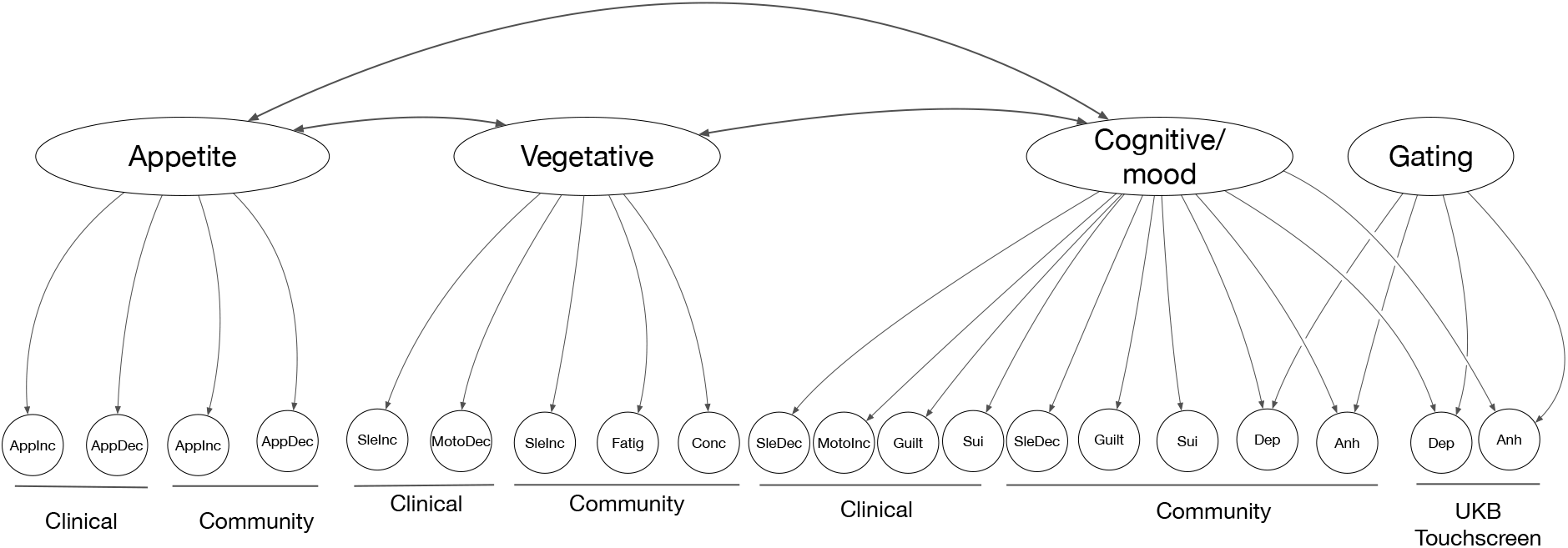
Model I: Appetite-Vegetative-Cognitive/Mood

**Figure S1j:**
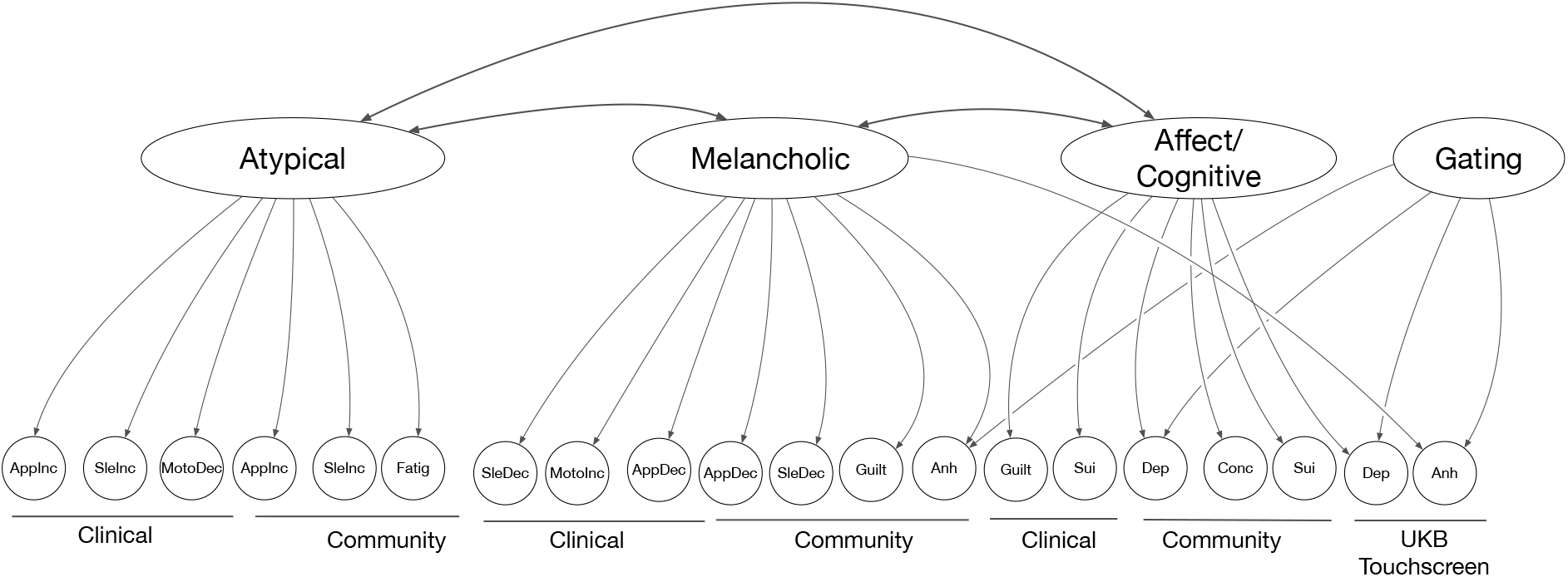
Model J: Atypical-Melancholic-Affect/Cognitive

### Symptom genetic correlations

**Supplementary Figure S2.**
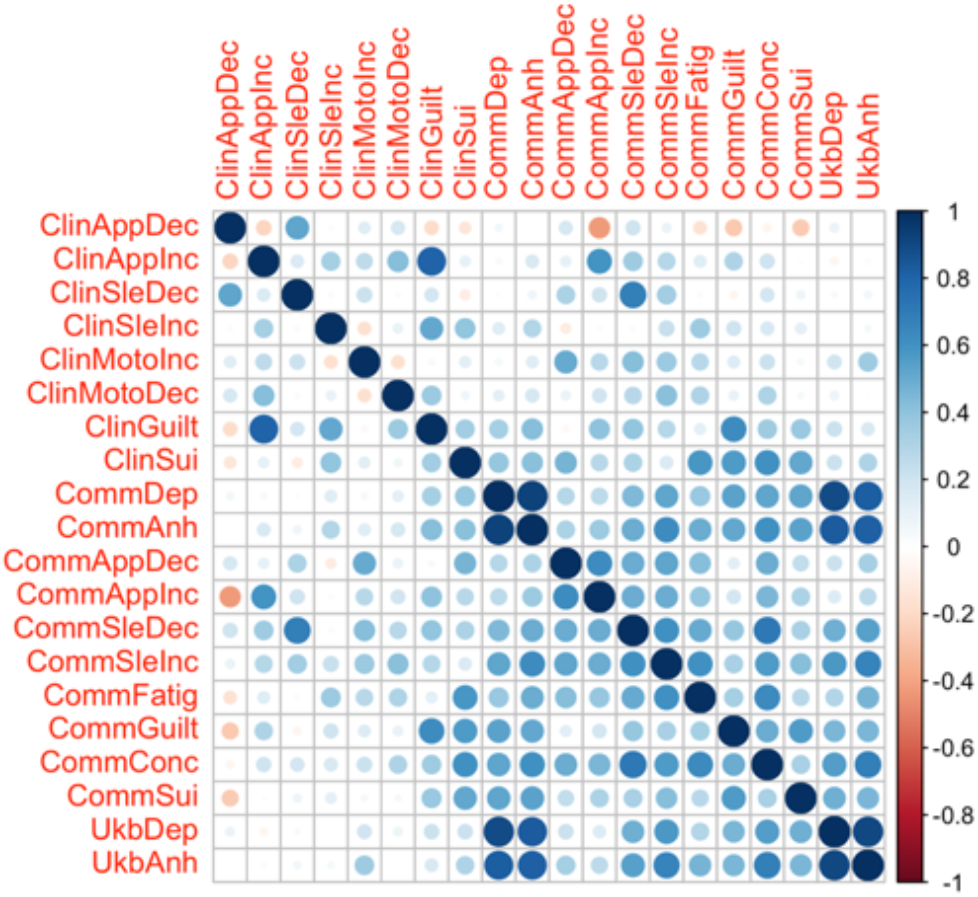
Genetic correlations between symptoms

Supplementary Figure S3. Model implied and residual proportions of genetic correlations

Variance and covariances scaled by total genetic variance of each symptom.

**Supplementary Figure S3a.**
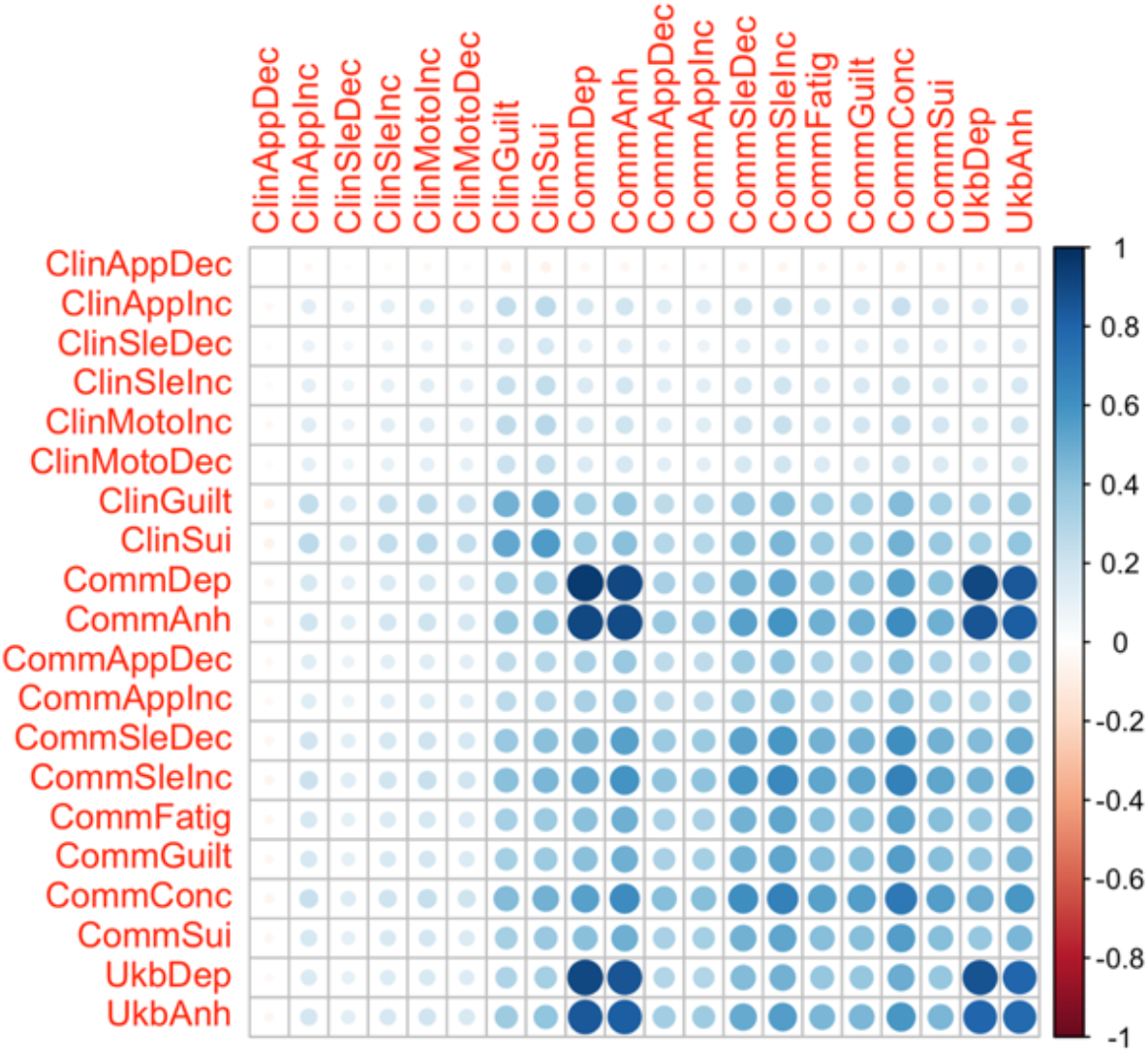
Model implied proportions of genetic correlations for Clinical Community-Gating factors (Model D)

**Supplementary Figure S3b.**
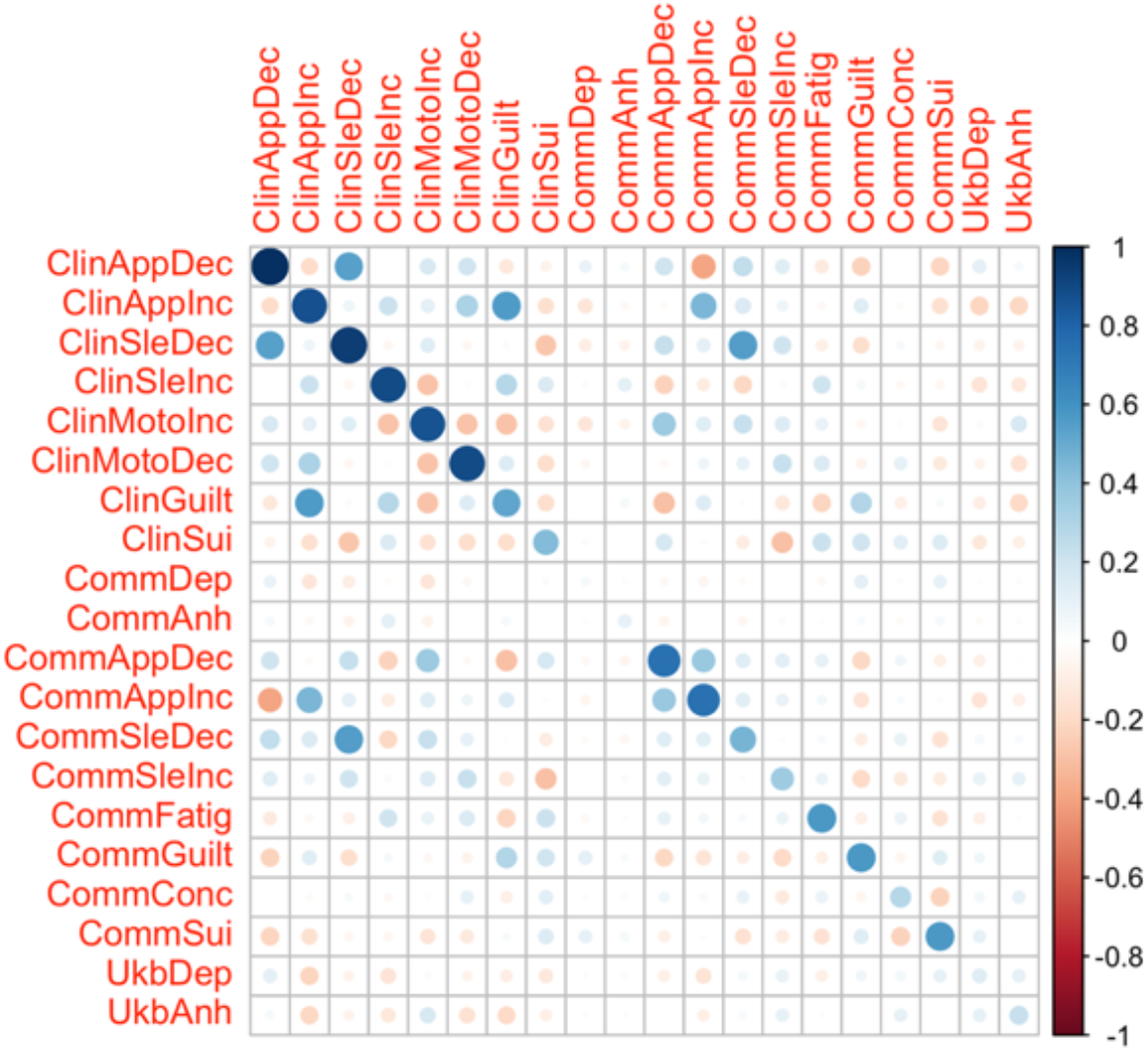
Model residual proportions of genetic correlations for Clinical Community-Gating factors (Model D)

**Supplementary Figure S3c.**
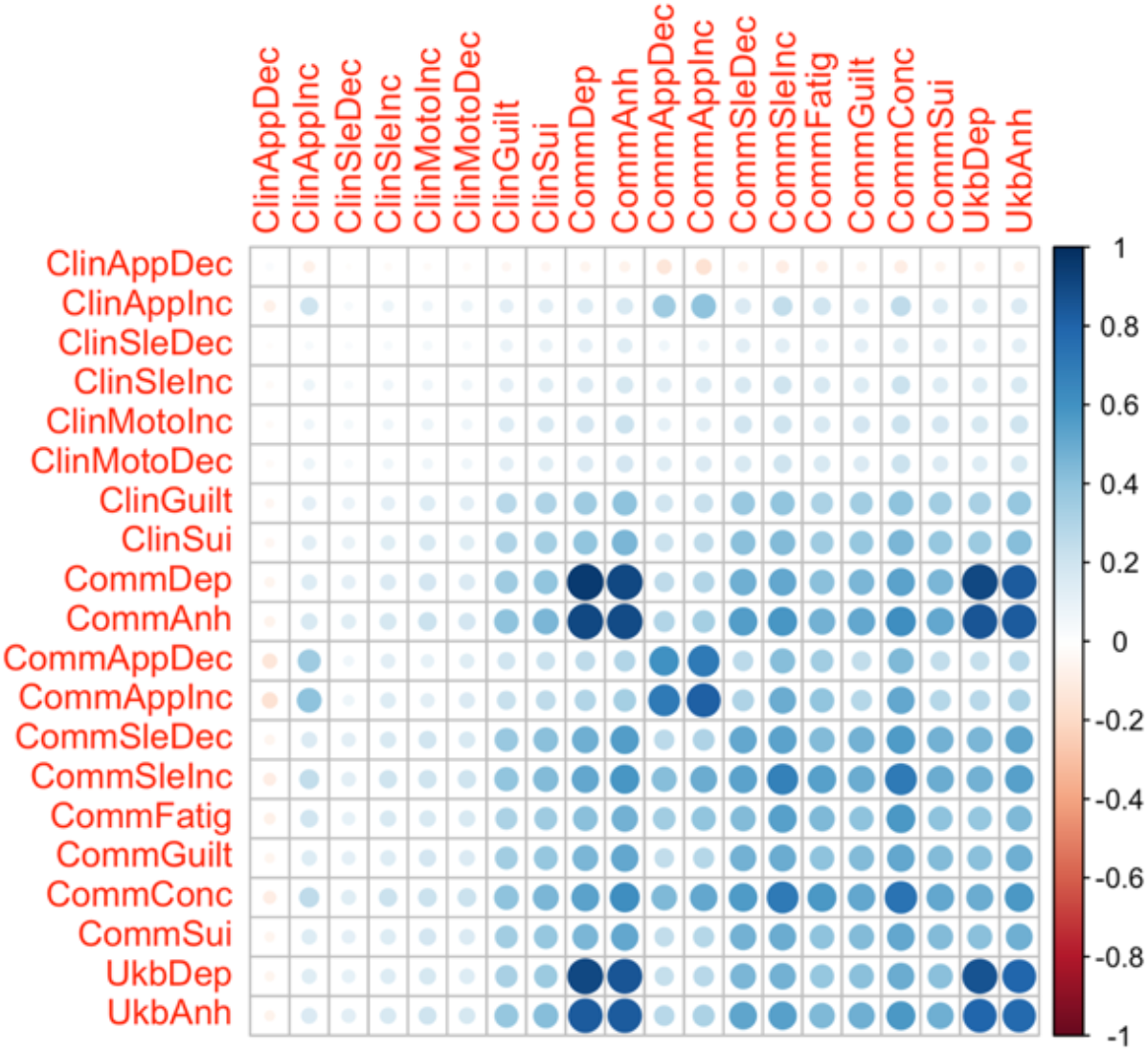
Model implied proportions of genetic correlations for Appetite Vegetative-Cognitive/Mood factors (Model I)

**Supplementary Figure S3d.**
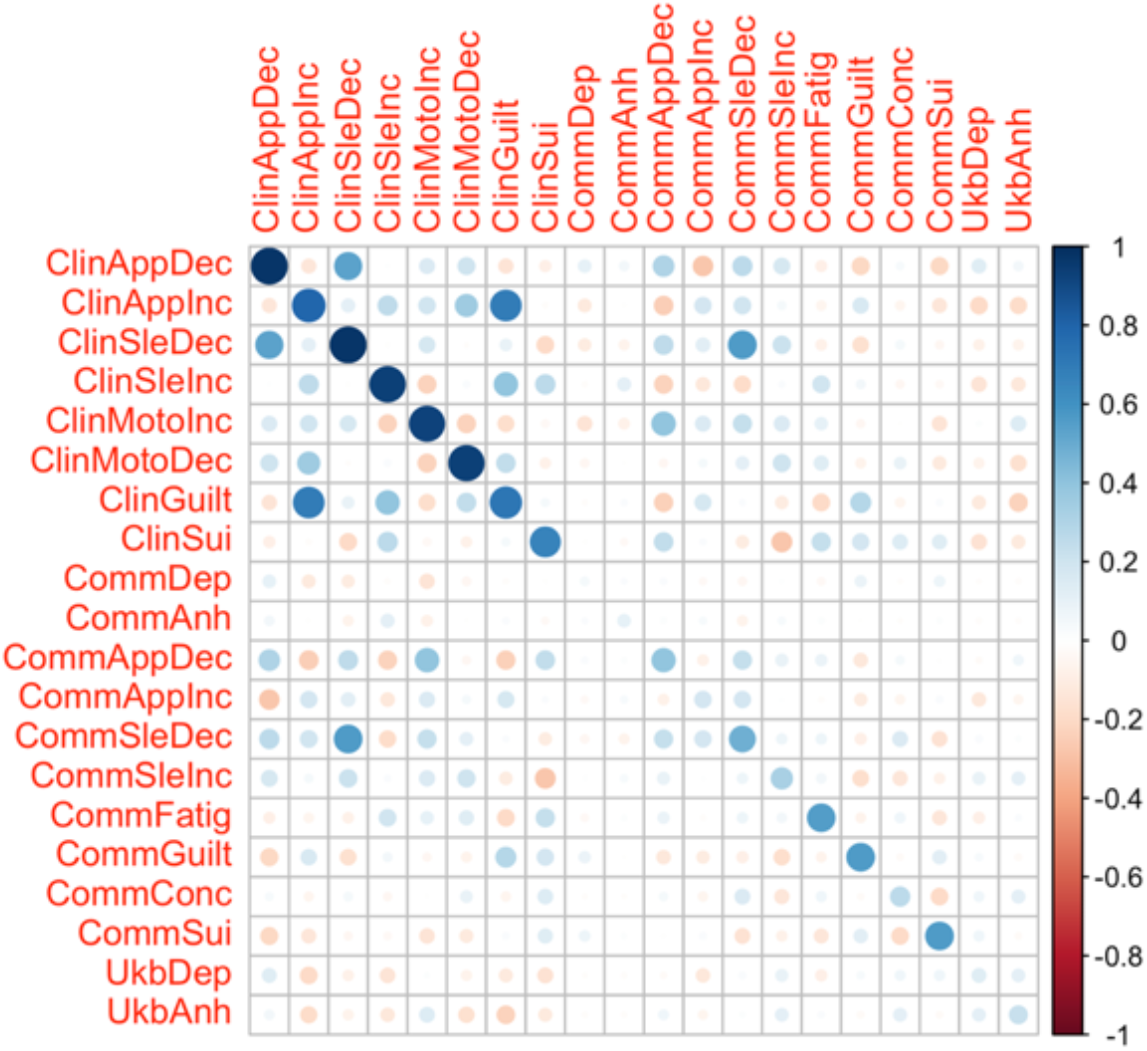
Model residual proportions of genetic correlations for Appetite Vegetative-Cognitive/Mood factors (Model I)

### External phenotype summary statistics

For the genetic multiple regression analysis, we used the following summary statistics:

- Alcohol dependence (Walters et al., 2018)
- Anxiety (Grotzinger et al., 2022)
- Bipolar disorder (Mullins et al., 2021)
- Body mass index (Pulit et al., 2019)
- Educational attainment (Okbay et al., 2022)
- Major depression (Als et al., 2022)
- Major depressive disorder (Wray et al., 2018)
- Neuroticism (Nagel et al., 2018)
- Pain (multisite chronic pain) (Johnston et al., 2019)
- Post-traumatic stress disorder (Nievergelt et al., 2019)
- Sleep (long sleep duration) (Dashti et al., 2018)
- Smoking (cigarettes per day) (Liu et al., 2019)

### Major Depressive Disorder Working Group of the Psychiatric Genomics Consortium

Mark J Adams 1

Fabian Streit 2

Swapnil Awasthi 3

Brett N Adey 4

Karmel W Choi 5, 6

V Kartik Chundru 7

Jonathan RI Coleman 4, 8

Jerome C Foo 2

Olga Giannakopoulou 9

Alisha S M Hall 2, 10

Jens Hjerling-Leffler 11

David M Howard 4

Christopher Hübel 4, 12, 13

Alex S F Kwong 1, 14

Bochao Danae Lin 15

Xiangrui Meng 9

Guiyan Ni 16

Oliver Pain 17

Gita A Pathak 18, 19

Eva C Schulte 20, 21, 22, 23

Jackson G Thorp 24

Alicia Walker 16

Shuyang Yao 25

Jian Zeng 16

Johan Zvrskovec 4, 8

Dag Aarsland 26

Ky’Era V Actkins 27

Mazda Adli 3, 28

Esben Agerbo 12, 29, 30

Mareike Aichholzer 31

Tracy M Air 32

Allison Aiello 33

Thomas D Als 30, 34, 35

Evelyn Andersson 36

Till F M Andlauer 37, 38

Volker Arolt 39

Helga Ask 40, 41

Sunita Badola 42

Clive Ballard 43

Karina Banasik 44

Nicholas J Bass 9

Aartjan T F Beekman 45

Sintia Belangero 46

Elisabeth B Binder 38, 47

Ottar Bjerkeset 48, 49

Gyda Bjornsdottir 50

Julia Boberg 36

Sigrid Børte 51, 52, 53

Emma Bränn 54

Alice Braun 55

Thorsten Brodersen 56

Søren Brunak 44

Mie T Bruun 57

Pichit Buspavanich 58, 59

Jonas Bybjerg-Grauholm 60, 61

Enda M Byrne 62

Archie Campbell 63, 64

Megan L. Campbell 65

Enrique Castelao 66

Jorge Cervilla 67, 68

Boris Chaumette 69

Chia-Yen Chen 70

Zhengming Chen 71, 72

Sven Cichon 73, 74, 75, 76

Lucía Colodro-Conde 24

Anne Corbett 43

Elizabeth C Corfield 40, 77

Baptiste Couvy-Duchesne 78

Nick Craddock 79, 80

Udo Dannlowski 39

Gail Davies 81

EJC de Geus 82

Ian J Deary 81

Franziska Degenhardt 76, 83

Abbas Dehghan 84, 85

J Raymond DePaulo 86

Michael Deuschle 87

Maria Didriksen 88

Khoa Manh Dinh 89

Nese Direk 90

Srdjan Djurovic 91, 92

Anna R Docherty 93, 94, 95

Katharina Domschke 96

Joseph Dowsett 88

Ole Kristian Drange 49, 97, 98, 99

Erin C Dunn 6, 100

Gudmundur Einarsson 50

Thalia C Eley 4

Samar S M Elsheikh 101

Jan Engelmann 102

Michael E Benros 60, 103, 104

Christian Erikstrup 89

Valentina Escott-Price 80

Chiara Fabbri 4, 105

Yu Fang 106

Sarah Finer 107

Josef Frank 2

Robert C Free 108

He Gao 109

Michael Gill 110

Maria Gilles 87

Fernando S Goes 86

Scott Douglas Gordon 24

Jakob Grove 30, 34, 35, 111

Daniel F Gudbjartsson 50, 112

Blanca Gutierrez 67, 68

Tim Hahn 39

Lynsey S Hall 80

Thomas F Hansen 44, 60, 113

Magnus Haraldsson 114

Catherina A Hartman 115

Alexandra Havdahl 40

Caroline Hayward 116

Stefanie Heilmann-Heimbach 76

Stefan Herms 74, 76

Ian B Hickie 117

Henrik Hjalgrim 118

Per Hoffmann 74, 76

Georg Homuth 119

Carsten Horn 120

Jouke-Jan Hottenga 82

David M Hougaard 60, 61

Iiris Hovatta 121

Qin Qin Huang 7

Floris Huider 82

Karen A Hunt 122

Marcus Ising 123

Erkki Isometsä 124

Rick Jansen 45

Yunxuan Jiang 125

Ian Jones 80

Lisa A Jones 126

Lina Jonsson 127

Robert Karlsson 25

Siegfried Kasper 128

Kenneth S Kendler 129

Ronald C Kessler 130

Stefan Kloiber 101, 123, 131, 132

James A Knowles 133

Nastassja Koen 65

Julia Kraft 55

Henry R Kranzler 134, 135

Kristi Krebs 136

Theodora Kunovac Kallak 137

Zoltán Kutalik 138, 139, 140

Elisa Lahtela 141

Margit Hørup Larsen 88

Eric J Lenze 142

Daniel F Levey 143, 144

Melissa Lewins 1

Glyn Lewis 9

Liming Li 145, 146

Kuang Lin 71

Penelope A Lind 24

Donald J MacIntyre 1, 147, 148

Dean F MacKinnon 86

Wolfgang Maier 149

Victoria S Marshe 101, 150

Hamdi Mbarek 82

Peter McGuffin 4

Sarah E Medland 24

Susanne Meinert 39, 151

Susan Mikkelsen 89

Christina Mikkelsen 88, 152

Yuri Milaneschi 45

Iona Y Millwood 71, 72

Brittany L Mitchell 24

Esther Molina 67, 153

Francis M Mondimore 86

Preben Bo Mortensen 12, 29, 30

Benoit H Mulsant 101, 131

Joonas Naamanka 121

Jake M Najman 154

Matthias Nauck 155, 156

Igor Nenadi? 157

Kasper R Nielsen 158

Ilja M Nolte 159

Merete Nordentoft 60, 103, 104

Markus M Nöthen 76

Mette Nyegaard 30, 160, 161, 162

Michael C O’Donovan 80

Asmundur Oddsson 50

Catherine M Olsen 163, 164

Hogni Oskarsson 165

Sisse Rye Ostrowski 88, 166

Vanessa K Ota 46

Michael J Owen 80

Richard Packer 167

Teemu Palviainen 141

Pedro M Pan 168

Carlos N Pato 169

Michele T Pato 169

Nancy L Pedersen 25

Ole Birger Pedersen 170

Roseann E Peterson 129, 171

Wouter J Peyrot 45

James B Potash 86

Martin Preisig 66

Jorge A Quiroz 172

Charles F Reynolds III 173

John P Rice 142

Giovanni A Salum 174

Robert A Schoevers 175, 176

Andrew Schork 30, 177, 178

Thomas G Schulze 2, 21, 86, 179, 180

Tabea S Send 87

Jianxin Shi 181

Engilbert Sigurdsson 114

Kritika Singh 27

Grant C B Sinnamon 182

Lea Sirignano 2

Olav B Smeland 183, 184

Daniel J Smith 185

Erik Sørensen 88

Sundararajan Srinivasan 186

Hreinn Stefansson 50

Kari Stefansson 50, 187

Dan J. Stein 188

Frederike Stein 189

André Tadic 102, 190

Henning Teismann 191

Alexander Teumer 192

Anita Thapar 80, 193

Pippa A Thomson 64

Lise Wegner Thørner 88

Apostolia Topaloudi 194

Ioanna Tzoulaki 84, 85, 195

Monica Uddin 196

André G Uitterlinden 197

Henrik Ullum 88, 198, 199

Daniel Umbricht 200

Robert J Ursano 201

Sandra Van der Auwera 202

David A van Heel 122

Albert M van Hemert 203

Abirami Veluchamy 186

Alexander Viktorin 25

Henry Völzke 192

Agaz Wani 196

G Bragi Walters 50

Robin G Walters 71, 72

Sylvia Wassertheil-Smoller 204

Myrna M Weissman 205, 206

Jürgen Wellmann 191

David C Whiteman 163

Derek Wildman 196

Gonneke Willemsen 82

Alexander T Williams 167

Bendik S Winsvold 51, 52, 207

Stephanie H Witt 2

Ying Xiong 25

Lea Zillich 2

John-Anker Zwart 51, 52, 53

23andMe Research Team 125

Estonian Biobank Research Team 136

HUNT All-In Psychiatry 208

China Kadoorie Biobank Collaborative Group 209

Genes & Health Research Team 210

Ole A Andreassen 183, 184, 211

Bernhard T Baune 212, 213, 214

Klaus Berger 191

Dorret I Boomsma 82

Anders D Børglum 30, 34, 35

Gerome Breen 4, 8

Na Cai 215, 216, 217

Hilary Coon 94

William E Copeland 218

Byron Creese 43

Lea K Davis 27

Eske M Derks 24

Enrico Domenici 219

Paul Elliott 84, 85, 195, 220

Andreas J Forstner 73, 76

Micha Gawlik 221

Joel Gelernter 19, 143, 222

Hans J Grabe 202

Steven P Hamilton 223

Kristian Hveem 224, 225, 226

Catherine John 167, 227

Jaakko Kaprio 141

Tilo Kircher 157

Marie-Odile Krebs 228

Karoline Kuchenbaecker 9, 71

Mikael Landén 25, 127

Kelli Lehto 136

Douglas F Levinson 229

Qingqin S Li 230

Klaus Lieb 102

Yi Lu 25

Susanne Lucae 123

Jurjen J Luykx 15, 231

Patrik K Magnusson 25

Nicholas G Martin 24

Hilary C Martin 7

Andrew McQuillin 9

Christel M Middeldorp 62, 232

Lili Milani 136

Ole Mors 30, 233

Daniel J Müller 101, 131, 132, 234

Bertram Müller-Myhsok 38, 235, 236

Albertine J Oldehinkel 115

Sara A Paciga 237

Colin NA Palmer 186

Peristera Paschou 194

Brenda WJH Penninx 45

Roy H Perlis 5, 6, 238

Giorgio Pistis 66

Renato Polimanti 18, 19

David J Porteous 64

Danielle Posthuma 239, 240

Ted Reichborn-Kjennerud 40

Andreas Reif 31

Frances Rice 80, 241

Roland Ricken 3

Marcella Rietschel 2

Margarita Rivera 67, 242

Christian Rück 243

Catherine Schaefer 244

Srijan Sen 106, 245

Alessandro Serretti 105

Alkistis Skalkidou 137

Jordan W Smoller 5, 246, 247

Frederike Stein 189

Murray B Stein 248, 249, 250

Patrick F Sullivan 25, 251

Martin Tesli 40

Thorgeir E Thorgeirsson 50

Henning Tiemeier 252, 253

Nicholas J Timpson 14

Rudolf Uher 254

Jens R Wendland 42

Thomas Werge 60, 177, 199, 255, 256

Naomi R Wray 16, 257

Stephan Ripke 3, 246

Cathryn M Lewis 4, 258

Andrew M McIntosh 1, 25

1. Division of Psychiatry, University of Edinburgh, Edinburgh, UK
2. Department of Genetic Epidemiology in Psychiatry, Central Institute of Mental Health, Medical Faculty Mannheim, Heidelberg University, Mannheim, BW, DE
3. Department of Psychiatry and Psychotherapy, Charité – Universitätsmedizin Berlin, Berlin, BE, DE
4. Social, Genetic and Developmental Psychiatry Centre, King’s College London, London, UK
5. Department of Psychiatry, Massachusetts General Hospital, Boston, MA, US
6. Department of Psychiatry, Harvard Medical School, Boston, MA, US
7. Human Genetics, Wellcome Sanger Institute, Hinxton, UK
8. NIHR Maudsley Biomedical Research Centre, King’s College London, London, UK
9. Division of Psychiatry, University College London, London, UK
10. Department of Clinical Medicine, Aarhus University, Aarhus, DK
11. Department of Medical Biochemistry and Biophysics, Karolinska Institutet, Stockholm, SE
12. National Centre for Register-based Research, Aarhus University, Aarhus, DK
13. Department of Pediatric Neurology, Charité – Universitätsmedizin Berlin, Berlin, BE, DE
14. MRC Integrative Epidemiology Unit, University of Bristol, Bristol, UK
15. Department of Psychiatry and Neuropsychology, School for Mental Health and Neuroscience, Maastricht University Medical Centre, Maastricht, NL
16. Institute for Molecular Bioscience, University of Queensland, Brisbane, QLD, AU
17. Maurice Wohl Clinical Neuroscience Institute, Department of Basic and Clinical Neuroscience, King’s College London, London, UK
18. Veterans Affairs Connecticut Healthcare System, West Haven, CT, US
19. Department of Psychiatry, Yale University School of Medicine, New Haven, CT, US
20. Department of Psychiatry, University of Munich, Munich, BY, DE
21. Institute of Psychiatric Phenomics and Genomics, University of Munich, Munich, BY, DE
22. Department of Psychiatry and Psychotherapy, University Hospital Bonn, Medical Faculty, University of Bonn, Bonn, DE
23. Institute of Human Genetics, University Hospital Bonn, Medical Faculty, University of Bonn, Bonn, DE
24. Mental Health and Neuroscience, QIMR Berghofer Medical Research Institute, Brisbane, QLD, AU
25. Department of Medical Epidemiology and Biostatistics, Karolinska Institutet, Stockholm, SE
26. Old Age Psychiatry, King’s College London, London, UK
27. Department of Medicine, Division of Genetic Medicine, Vanderbilt University Medical Center, Nashville, TN, US
28. Department of Psychiatry and Psychotherapy, Fliedner Klinik Berlin, Berlin, BE, DE
29. Centre for Integrated Register-based Research, Aarhus University, Aarhus, DK
30. iPSYCH, The Lundbeck Foundation Initiative for Integrative Psychiatric Research, Aarhus, DK
31. Department of Psychiatry, Psychosomatic Medicine and Psychotherapy, Goethe University Frankfurt - University Hospital, Frankfurt am Main, DE
32. Discipline of Psychiatry, University of Adelaide, Adelaide, SA, AU
33. Department of Epidemiology, Columbia University Mailman School of Public Health, New York, NY, US
34. Department of Biomedicine and Centre for Integrative Sequencing, iSEQ, Aarhus University, Aarhus, DK
35. Center for Genomics and Personalized Medicine, Aarhus University, Aarhus, DK
36. Department of Clinical Neuroscience, Karolinska Institutet,, SE
37. Department of Neurology, Klinikum rechts der Isar, Technical University of Munich, Munich, BY, DE
38. Department of Translational Research in Psychiatry, Max Planck Institute of Psychiatry, Munich, BY, DE
39. Institute for Translational Psychiatry, University of Münster, Münster, NRW, DE
40. Department of Mental Disorders, Norwegian Institute of Public Health, Oslo, NO
41. PROMENTA Research Center, Department of Psychology, University of Oslo, Oslo, NO
42. Research and Development, Takeda Pharmaceutical Company Limited, Cambridge, MA, US
43. Faculty of Health and Life Sciences, University of Exeter, Exeter, UK
44. Novo Nordisk Center for Protein Research, Department of Health Sciences, University of Copenhagen, Copenhagen, DK
45. Department of Psychiatry, Amsterdam Public Health and Amsterdam Neuroscience, Amsterdam UMC, Vrije Universiteit Amsterdam, Amsterdam, NL
46. Morphology and Genetics, Universidade Federal de Sao Paulo, Sao Paulo, SP, BR
47. Department of Psychiatry and Behavioral Sciences, Emory University School of Medicine, Atlanta, GA, US
48. Faculty of Nursing and Health Sciences, NORD University, Levanger, NO
49. Department of Mental Health, Faculty of Medicine and Health Sciences, Norwegian University of Science and Technology (NTNU), Trondheim, TRD, NO
50. deCODE Genetics / Amgen, Reykjavik, IS
51. K. G. Jebsen Center for Genetic Epidemiology, Department of Public Health and Nursing, Faculty of Medicine and Health Sciences, Norwegian University of Science and Technology (NTNU), Trondheim, TRD, NO
52. Department of Research and Innovation, Division of Clinical Neuroscience, Oslo University Hospital, Oslo, NO
53. Institute of Clinical Medicine, Faculty of Medicine, University of Oslo, Oslo, NO
54. Institute of Environmental Medicine, Unit of Integrative Epidemiology, Karolinska Institutet, Stockholm, SE
55. Department of Psychiatry and Psychotherapy, Charité – Universitätsmedizin Berlin, Berlin, DE
56. Department of Clinical Immunology, Roskilde University/Næstved Hospital, Roskilde, DK
57. Department of Clinical Immunology, Odense University Hospital, Odense, DK
58. Department of Psychiatry, Psychotherapy and Psychosomatics, Brandenburg Medical School Theodor Fontane, Neuruppin, BB, DE
59. Department of Psychiatry and Psychotherapy, Gender Research in Medicine, Institute of Sexology and Sexual Medicine, Charité – Universitätsmedizin Berlin, Berlin, BE, DE
60. iPSYCH, The Lundbeck Foundation Initiative for Integrative Psychiatric Research, Copenhagen, DK
61. Center for Neonatal Screening, Department for Congenital Disorders, Statens Serum Institut, Copenhagen, DK
62. Child Health Research Centre, University of Queensland, Brisbane, QLD, AU
63. Centre for Medical Informatics, Usher Institute, University of Edinburgh, Edinburgh, UK
64. Centre for Genomic & Experimental Medicine, Institute for Genetics and Cancer, University of Edinburgh, Edinburgh, UK
65. Department of Psychiatry and Mental Health, University of Cape Town, Cape Town, SA
66. Department of Psychiatry, Lausanne University Hospital and University of Lausanne, Prilly, VD, CH
67. Instituto de Investigación Biosanitaria ibs.GRANADA, Granada, ES
68. Department of Psychiatry, Faculty of Medicine and Institute of Neurosciences, Biomedical Research Centre (CIBM), University of Granada, Granada, ES
69. Université de Paris Cité, INSERM U1266, Institute of Psychiatry and Neuroscience of Paris, GHU Paris Psychiatry and Neuroscience, Paris, FR
70. Translational Biology, Biogen, Cambridge, MA, US
71. Nuffield Department of Population Health, University of Oxford, Oxford, UK
72. MRC Population Health Research Unit, University of Oxford, Oxford, UK
73. Institute of Neuroscience and Medicine (INM-1), Research Center Juelich, Juelich, DE
74. Human Genomics Research Group, Department of Biomedicine, University of Basel, Basel, CH
75. Institute of Medical Genetics and Pathology, University Hospital Basel, University of Basel, Basel, CH
76. Institute of Human Genetics, University of Bonn, School of Medicine & University Hospital Bonn, Bonn, DE
77. Nic Waals Institute, Lovisenberg Diakonale Hospital, Oslo, NO
78. Centre for Advanced Imaging, University of Queensland, Saint Lucia, QLD, AU
79. Psychological Medicine, Cardiff University, Cardiff, WLS, UK
80. Centre for Neuropsychiatric Genetics and Genomics, Cardiff University, Cardiff, WLS, UK
81. The Lothian Birth Cohorts, University of Edinburgh, Edinburgh, UK
82. Department of Biological Psychology & Amsterdam Public Health Research Institute, Vrije Universiteit Amsterdam, Amsterdam, NL
83. Department of Child and Adolescent Psychiatry, Psychosomatics and Psychotherapy, University Hospital Essen, Unversity of Duisburg-Essen, Duisburg, DE
84. MRC Centre for Environment and Health, School of Public Health, Imperial College London, London, UK
85. Imperial College Dementia Research Institute, Imperial College London, London, UK
86. Department of Psychiatry and Behavioral Sciences, Johns Hopkins University School of Medicine, Baltimore, MD, US
87. Department of Psychiatry and Psychotherapy, Research Group Stress Related Disorders, Central Institute of Mental Health, Medical Faculty Mannheim, Heidelberg University, Mannheim, BW, DE
88. Department of Clinical Immunology, Copenhagen University Hospital, Rigshospitalet, Copenhagen, CPH, DK
89. Department of Clinical Immunology, Aarhus University Hospital, Aarhus, DK
90. Department of Psychiatry, Istanbul University, Istanbul, TR
91. Department of Medical Genetics, Oslo University Hospital, Oslo, OSL, NO
92. NORMENT, Department of Clinical Science, University of Bergen, Bergen, NO
93. Virginia Institute for Psychiatric & Behavioral Genetics, Virginia Commonwealth University, Richmond, VA, US
94. Psychiatry Department / Huntsman Mental Health Institute, University of Utah School of Medicine, Salt Lake City, UT, US
95. Center for Genomic Research, University of Utah School of Medicine, Salt Lake City, UT, US
96. Department of Psychiatry and Psychotherapy, Medical Center, University of Freiburg, Faculty of Medicine, University of Freiburg, Freiburg, DE
97. Division of Mental Health Care, St. Olavs Hospital, Trondheim University Hospital, Trondheim, TRD, NO
98. Department of Psychiatry, Sørlandet Hospital, Kristiansand, AG, NO
99. University of Oslo, NORMENT Centre, Institute of Clinical Medicine, Oslo, OSL, NO
100. Center for Genomic Medicine, Massachusetts General Hospital, Boston, MA, US
101. Centre for Addiction and Mental Health, Toronto, ON, CA
102. Department of Psychiatry and Psychotherapy, University Medical Center of the Johannes Gutenberg University Mainz, Mainz, DE
103. Mental Health Center Copenhagen, Mental Health Services Capital Region of Denmark, Copenhagen, DK
104. Faculty of Health Science, Department of Clinical Medicine, University of Copenhagen, Copenhagen, DK
105. Department of Biomedical and Neuromotor Sciences, University of Bologna, Bologna, IT
106. Michigan Neuroscience Institute, University of Michigan, Ann Arbor, MI, US
107. Wolfson Institute of Population Health, Queen Mary University of London, London, UK
108. School of Computing and Mathematical Sciences, University of Leicester, Leicester, UK
109. Department of Epidemiology and Biostatistics, Imperial College London, London, UK
110. Discipline of Psychiatry, School of Medicine, Trinity College Dublin, Dublin, IE
111. Bioinformatics Research Centre, Aarhus University, Aarhus, DK
112. 112, School of Engineering, University of Iceland, Reykjavik, IS
113. Danish Headache Centre, Department of Neurology, Rigshospitalet, Glostrup, DK
114. Faculty of Medicine, Department of Psychiatry, University of Iceland, Reykjavik, IS
115. Department of Psychiatry, University of Groningen, University Medical Center Groningen, Groningen, NL
116. MRC Human Genetics Unit, Institute for Genetics and Cancer, University of Edinburgh, Edinburgh, UK
117. Brain and Mind Centre, University of Sydney, Sydney, NSW, AU
118. Department of Epidemiology Research, Statens Serum Institut, Copenhagen, DK
119. Interfaculty Institute for Genetics and Functional Genomics, Department of Functional Genomics, University Medicine Greifswald, Greifswald, MV, DE
120. Roche Pharmaceutical Research and Early Development, Pharmaceutical Sciences, Roche Innovation Center Basel, F. Hoffmann-La Roche Ltd, Basel, CH
121. SleepWell Research Program and Department of Psychology and Logopedics, University of Helsinki, Helsinki, FI
122. Blizard Institute, Barts and the London School of Medicine and Dentistry, Queen Mary University of London, London, UK
123. Max Planck Institute of Psychiatry, Munich, BY, DE
124. Department of Psychiatry, University of Helsinki, Helsinki, FI
125. 23andMe Research Team, 23andMe, Inc., Sunnyvale, CA, US
126. Department of Psychological Medicine, University of Worcester, Worcester, UK
127. Institution of Neuroscience and Physiology, University of Gothenburg, Gothenburg, SE
128. Department of Psychiatry and Psychotherapy, Medical University of Vienna, Vienna, AT
129. Department of Psychiatry, Virginia Commonwealth University, Richmond, VA, US
130. Health Care Policy, Harvard Medical School, Boston, MA, US
131. Department of Psychiatry, University of Toronto, Toronto, ON, CA
132. Department of Pharmacology & Toxicology, University of Toronto, Toronto, ON, CA
133. Department of Genetics, Rutgers University, Piscataway, NJ, US
134. Department of Psychiatry, Perelman School of Medicine, University of Pennsylvania, Philadelphia, PA, US
135. Mental Illness Research, Education and Clinical Center, Crescenz VA Medical Center, Philadelphia, PA, US
136. Estonian Genome Centre, Institute of Genomics, University of Tartu, Tartu, EE
137. Department of Women’s and Children’s Health, Uppsala University, Uppsala, SE
138. Department of Epidemiology and Health Systems, Center for Primary Care and Public Health, Lausanne, VD, CH
139. Swiss Institute of Bioinformatics, Lausanne, VD, CH
140. Department of Computational Biology, University of Lausanne, Lausanne, VD, CH
141. Institute for Molecular Medicine Finland - FIMM, University of Helsinki, Helsinki, FI
142. Department of Psychiatry, Washington University School of Medicine in St. Louis, St. Louis, MO, US
143. Psychiatry, Veterans Affairs Connecticut Healthcare System, West Haven, CT, US
144. Department of Psychiatry, Yale University, New Haven, CT, US
145. Department of Epidemiology and Biostatistics, School of Public Health, Peking University, Beijing, CN
146. Peking University Center for Public Health and Epidemic Preparedness & Response, Peking University, Beijing, CN
147. Mental Health, NHS 24, Glasgow, UK
148. Royal Edinburgh Hospital, NHS Lothian, Edinburgh, UK
149. Department of Psychiatry and Psychotherapy, University of Bonn, Bonn, DE
150. Center for Translational and Computational Neuroimmunology, Columbia University Medical Center, New York, NY, US
151. Institute for Translational Neuroscience, University of Münster, Münster, NRW, DE
152. Novo Nordisk Foundation Center for Basic Metabolic Research, Faculty of Health Science, Copenhagen University, Copenhagen, DK
153. Department of Nursing, Faculty of Health Sciences and Institute of Neurosciences, Biomedical Research Centre (CIBM), University of Granada, Granada, ES
154. School of Public Health, University of Queensland, Brisbane, QLD, AU
155. DZHK (German Centre for Cardiovascular Research), Partner Site Greifswald, Greifswald, MV, DE
156. Institute of Clinical Chemistry and Laboratory Medicine, University Medicine Greifswald, Greifswald, MV, DE
157. Department of Psychiatry, University of Marburg, Marburg, DE
158. Department of Clinical Immunology, Aalborg University Hospital, Aalborg, DK
159. Department of Epidemiology, University of Groningen, University Medical Center Groningen, Groningen, NL
160. Department of Health, Science and Technology, Aalborg University, Aalborg, DK
161. Centre for Integrative Sequencing, iSEQ, Aarhus University, Aarhus, DK
162. Department of Biomedicine-Human Genetics, Aarhus University, Aarhus, DK
163. Population Health, QIMR Berghofer Medical Research Institute, Brisbane, QLD, AU
164. The Fraser Institute, Faculty of Medicine, University of Queensland, Brisbane, QLD, AU
165. Humus, Reykjavik, IS
166. Department of Clinical Medicine, University of Copenhagen, Copenhagen, CPH, DK
167. Department of Population Health Sciences, University of Leicester, Leicester, UK
168. Department of Psychiatry, Universidade Federal de Sao Paulo, Sao Paulo, SP, BR
169. Department of Psychiatry, Rutgers University, Piscataway, NJ, US
170. Department of Clinical Immunology, Zealand University Hospital, Køge, DK
171. Department of Psychiatry and Behavioral Sciences, SUNY Downstate Health Sciences University, Brooklyn, NY, US
172. NMD Pharma, Lexington, MA, US
173. Psychiatry, University of Pittsburgh Medical Centre, Pittsburgh, PA, US
174. Psychiatry, Universidade Federal do Rio Grande do Sul, Porto Alegre, BR
175. Department of Psychiatry, University Medical Center Groningen, Groningen, NL
176. Research School of Behavioural and Cognitive Neurosciences (BCN), University of Groningen, Groningen, NL
177. Institute of Biological Psychiatry, Mental Health Center Sct. Hans, Mental Health Services Capital Region of Denmark, Copenhagen, DK
178. Neurogenomics Division, The Translational Genomics Research Institute (TGEN), Phoenix, AZ, US
179. Human Genetics Branch, NIMH Division of Intramural Research Programs, Bethesda, MD, US
180. Department of Psychiatry and Psychotherapy, University Medical Center Göttingen, Goettingen, NI, DE
181. Division of Cancer Epidemiology and Genetics, National Cancer Institute, Bethesda, MD, US
182. School of Medicine and Dentistry, James Cook University, Townsville, QLD, AU
183. Division of Mental Health and Addiction, Oslo University Hospital, Oslo, OSL, NO
184. NORMENT, Institute of Clinical Medicine, University of Oslo, Oslo, OSL, NO
185. Institute of Health and Wellbeing, University of Glasgow, Glasgow, UK
186. Division of Population Health and Genomics, Ninewells Hospital and School of Medicine, University of Dundee, Dundee, UK
187. Faculty of Medicine, University of Iceland, Reykjavik, IS
188. SAMRC Unit on Risk & Resilience in Mental Disorders, Department of Psychiatry and Mental Health, University of Cape Town, Cape Town, SA
189. Department of Psychiatry and Psychotherapy, University of Marburg, Marburg, HE, DE
190. Department of Psychiatry, Psychotherapy and Psychosomatics, Dr. Fontheim Mentale Gesundheit, Liebenburg, DE
191. Institute of Epidemiology and Social Medicine, University of Münster, Münster, NRW, DE
192. Institute for Community Medicine, University Medicine Greifswald, Greifswald, MV, DE
193. Wolfson Centre for Young People’s Mental Health, Division of Psychological Medicine and Clinical Neurosciences, Cardiff University, Cardiff, WLS, UK
194. Department of Biological Sciences, Purdue University, West Lafayette, IN, US
195. Imperial College BHF Centre for Research Excellence, Imperial College London, London, UK
196. Genomics Program, University of South Florida College of Public Health, Tampa, FL, US
197. Department of Internal Medicine, Erasmus University Medical Center Rotterdam, Rotterdam, NL
198. Management Section, Statens Serum Institut, Copenhagen, DK
199. Department of Clinical Medicine, University of Copenhagen, Copenhagen, DK
200. Xperimed LLC, Basel, CH
201. Psychiatry, USUHS, Bethesda, US
202. Department of Psychiatry and Psychotherapy, University Medicine Greifswald, Greifswald, MV, DE
203. Department of Psychiatry, Leiden University Medical Center, Leiden, NL
204. Department of Epidemiology and Population Health, Albert Einstein College of Medicine, Bronx, NY, US
205. Department of Psychiatry, Columbia University College of Physicians and Surgeons, New York, NY, US
206. Division of Epidemiology, New York State Psychiatric Institute, New York, NY, US
207. Department of Neurology, Oslo University Hospital, Oslo, NO
208. HUNT All-In Psychiatry
209. China Kadoorie Biobank Collaborative Group
210. Genes & Health Research Team
211. KG Jebsen Centre for Neurodevelopmental Research, University of Oslo, Oslo, OSL, NO
212. Department of Psychiatry, University of Melbourne, Melbourne, VIC, AU
213. Florey Institute of Neuroscience and Mental Health, University of Melbourne, Melbourne, VIC, AU
214. Department of Psychiatry, University of Münster, Münster, NRW, DE
215. Computational Health Centre, Helmholtz Zentrum München, Neuherberg, DE
216. School of Medicine, Technical University of Munich, Munich, BY, DE
217. Helmholtz Pioneer Campus, Helmholtz Zentrum München, Neuherberg, DE
218. Department of Psychiatry, University of Vermont, Burlington, VT, US
219. Department of Cellular, Computational and Integrative Biology, Università degli Studi di Trento, Trento, IT
220. Imperial College Biomedical Research Centre, Imperial College London, London, UK
221. Department of Psychiatry, Psychosomatics and Psychotherapy, Julius-Maximilians-Universität Würzburg, Würzburg, DE
222. Department of Genetics, Department of Neuroscience, Yale University School of Medicine, New Haven, CT, US
223. Psychiatry, Kaiser Permanente Northern California, San Francisco, CA, US
224. K. G. Jebsen Center for Genetic Epidemiology, Department of Public Health and Nursing, Faculty of Medicine and Health Sciences, Norwegian University of Science and Technology (NTNU), Trondheim, NO
225. HUNT Research Center, Department of Public Health and Nursing, Faculty of Medicine and Health Sciences, Norwegian University of Science and Technology (NTNU), Trondheim, NO
226. Department of Research, Innovation and Education, St. Olavs Hospital, Trondheim University Hospital, Trondheim, NO
227. NIHR Leicester Biomedical Research Centre, Glenfield Hospital, Leicester, UK
228. Pathophysiology of Psychiatric Diseases, INSERM, Univ Paris Cité, GHU Paris, Paris, FR
229. Department of Psychiatry & Behavioral Sciences, Stanford University, Stanford, CA, US
230. Neuroscience Therapeutic Area, Janssen Research and Development, LLC, Titusville, NJ, US
231. Second Opinion Outpatient Clinic, GGNet Mental Health, Warnsveld, NL
232. Child and Youth Mental Health Service, Children’s Health Queensland Hospital and Health Service, Brisbane, QLD, AU
233. Psychosis Research Unit, Aarhus University Hospital-Psychiatry, Aarhus, DK
234. Department of Psychiatry, Psychosomatics and Psychotherapy, University Hospital of Würzburg, Würzburg, DE
235. Munich Cluster for Systems Neurology (SyNergy), Munich, BY, DE
236. University of Liverpool, Liverpool, UK
237. Human Genetics and Computational Biomedicine, Pfizer Global Research and Development, Groton, CT, US
238. Centre for Quantitative Health, Massachusetts General Hospital, Boston, MA, US
239. Child and Adolescent Psychiatry, Amsterdam UMC, Vrije Universiteit Amsterdam, Amsterdam, NL
240. Complex Trait Genetics, Vrije Universiteit Amsterdam, Amsterdam, NL
241. Wolfson Centre for Young People’s Mental Health, Division of Psychological Medicine and Clinical Neurosciences, Cardiff University, Cardiff, UK
242. Department of Biochemistry and Molecular Biology II, Faculty of Pharmacy and Institute of Neurosciences, Biomedical Research Centre (CIBM), University of Granada, Granada, ES
243. Department of Clinical Neuroscience, Karolinska Institutet, Stockholm, SE
244. Division of Research, Kaiser Permanente Northern California, Oakland, CA, US
245. Department of Psychiatry, University of Michigan, Ann Arbor, MI, US
246. Stanley Center for Psychiatric Research, Broad Institute of MIT and Harvard, Cambridge, MA, US
247. Psychiatric and Neurodevelopmental Genetics Unit, Massachusetts General Hospital, Boston, MA, US
248. Psychiatry, UCSD School of Medicine, La Jolla, CA, US
249. Public Health, UCSD School of Public Health, La Jolla, CA, US
250. Psychiatry, Veterans Affairs San Diego Healthcare System, San Diego, CA, US
251. Departments of Genetics and Psychiatry, University of North Carolina at Chapel Hill, Chapel Hill, NC, US
252. Child and Adolescent Psychiatry, Erasmus University Medical Center Rotterdam, Rotterdam, NL
253. Social and Behavioral Science, Harvard T.H. Chan School of Public Health, Boston, MA, US
254. Psychiatry, Dalhousie University, Halifax, NS, CA
255. Institute of Biological Psychiatry, Mental Health Center Sct. Hans, Copenhagen University Hospital, Mental Health Services, Copenhagen, DK
256. GLOBE Institute, Lundbeck Foundation Centre for Geogenetics, University of Copenhagen, Copenhagen, DK
257. Queensland Brain Institute, University of Queensland, Brisbane, QLD, AU
258. Department of Medical & Molecular Genetics, King’s College London, London, UK
259. Institute for Genomics and Cancer, University of Edinburgh, Edinburgh, UK

